# Functional genomics atlas of synovial fibroblasts defining rheumatoid arthritis heritability

**DOI:** 10.1101/2020.12.16.20248230

**Authors:** Xiangyu Ge, Mojca Frank-Bertoncelj, Kerstin Klein, Amanda Mcgovern, Tadeja Kuret, Miranda Houtman, Blaž Burja, Raphael Micheroli, Miriam Marks, Andrew Filer, Christopher D. Buckley, Gisela Orozco, Oliver Distler, Andrew P Morris, Paul Martin, Stephen Eyre, Caroline Ospelt

**Affiliations:** Versus Arthritis Centre for Genetics and Genomics, School of Biological Sciences, Faculty of Biology, Medicine and Health, The University of Manchester, Manchester, UK; Department of Rheumatology, Center of Experimental Rheumatology, University Hospital Zurich, University of Zurich, Zurich, Switzerland; Department of Rheumatology, University Medical Centre, Ljubljana, Slovenia; Schulthess Klinik, Zurich, Switzerland; Institute of Inflammation and Ageing, University of Birmingham, Birmingham, UK; NIHR Birmingham Biomedical Research Centre, University Hospitals Birmingham NHS Foundation Trust, University of Birmingham, Birmingham, UK; Kennedy Institute of Rheumatology, University of Oxford Roosevelt Drive Headington Oxford UK

## Abstract

Genome-wide association studies have reported >100 risk loci for rheumatoid arthritis (RA). These loci have been shown to be enriched in immune cell-specific enhancers, but analysis so far has excluded stromal cells, such as synovial fibroblasts (FLS), despite their crucial involvement in the pathogenesis of RA. Here we integrated DNA architecture (ChIP-seq), 3D chromatin interactions (HiC, capture HiC), DNA accessibility (ATAC-seq) and gene expression (RNA-seq) in FLS, B cells and T cells with genetic fine mapping of RA loci. We identified putative causal variants, enhancers, genes, and cell types for 30 - 60% of RA loci and demonstrated that FLS account for up to 24% of RA heritability. TNF stimulation of FLS altered the organization of topologically associating domains (TADs), chromatin state and the expression of putative causal genes (e.g. *TNFAIP3, IFNAR1)*. Several putative causal genes constituted RA-relevant functional networks in FLS with roles in cellular proliferation and activation. Finally, we demonstrated that risk variants can have joint-specific effects on target gene expression in RA FLS, which may contribute to the development of the characteristic pattern of joint involvement in RA. Overall, our research provides the first direct evidence for a causal role of FLS in the genetic susceptibility for RA accounting for up to a quarter of RA heritability.

## Main

A major challenge of the post-genome-wide association study (GWAS) era is to decipher the functional consequences of genetic risk variants in individual cell types and their contribution to the development of polygenic diseases. The identification of the cell types and conditions in which genetic risk variants are effective is an essential prerequisite for achieving this goal. Rheumatoid arthritis (RA) is a symmetric inflammatory and destructive autoimmune arthritis with a complex genetic basis. RA affects 0.5-1% of the world population and leads to disability, high morbidity burden and premature mortality^1^. GWAS have identified over 100 loci for RA susceptibility^2^. Genetic risk variants at the majority of these loci do not map to the exons of protein coding genes. Potential gene regulatory functions of these noncoding genetic risk variants have been investigated in immune cells based on genome-wide mapping of epigenetic modifications^3^, chromatin interactions^4^, correlation with variation in gene expression (eQTLs)^5^ or linear proximity to coding genes in DNA sequence^2^. These studies have demonstrated an enrichment of RA genetic risk variants in immune cell enhancers^3^, but omitted the analysis of synovial fibroblasts or Fibroblast-like Synoviocytes (FLS), the resident stromal cells of the joints, even though they are responsible for the production of many immune related cytokines and chemokines^6,7^.

In addition to immune cells, FLS play a decisive role in the pathogenesis of RA and are essential for the maintenance of normal joint functions. FLS from different joints have different epigenomes, transcriptomes and functions, which may contribute to the characteristic pattern of joint involvement in different types of arthritis^8,9^. FLS substantially contribute to joint inflammation and destruction in RA^10^. RA FLS have an activated phenotype characterized by resistance to apoptosis, increased proliferation, secretion of matrix-degrading enzymes and production of cytokines and chemokines that promote immune cell differentiation and survival. However, the cause of the activation of FLS in RA is unknown and it is unclear whether this activation leads to or is a consequence of the disease. Defining the contribution of FLS to the heritability of RA will provide essential insights into this question.

For the first time, we have comprehensively mapped RA genetic risk variants to active regulatory DNA elements in FLS. We generated multidimensional epigenetic data in primary FLS, isolated from patients, to create a detailed outline of their chromatin landscape. We conducted genetic fine-mapping of RA loci by computing sets of credible single nucleotide polymorphisms (SNPs) driving GWAS signals. We integrated the credible SNP sets and chromatin datasets to provide evidence that RA risk variants can be functionally relevant in FLS. We used chromatin conformation data to determine enhancer–promoter interactions between risk variants in non-coding DNA regulatory regions of FLS and their target genes. Furthermore, we assessed the influence of the pro-inflammatory cytokine tumour necrosis factor (TNF) on these interactions, chromatin accessibility and gene expression in FLS. We combined FLS data with published data of human tissues and cells^4,11,12^ to identify putative causal SNPs, enhancers, genes and cell types for RA risk loci. Finally, we functionally verified enhancer-promoter interactions by CRISPR-Cas technology and showed transcriptional effects of fine-mapped risk variants in FLS samples from RA patients.

## Results

### Integration of epigenetic datasets to define the chromatin landscape of FLS

As a first step in our analysis, we generated diverse epigenetic and transcriptomic datasets from our primary FLS samples (Supplementary Table 1): chromatin immunoprecipitation sequencing (ChIP-seq) for six histone marks (H3K4me3, H3K4me1, H3K27me3, H3K36me3, H3K27ac, H3K9me3), Assay for Transposase-Accessible Chromatin sequencing (ATAC-seq), cap analysis gene expression sequencing (CAGE-seq), chromatin conformation analysis (HiC, Capture HiC) and RNA sequencing (RNA-seq) (Supplementary Table 2, quality control metrics in Supplementary Dataset 1, details in Online methods). We used these datasets to annotate open chromatin and to assign 18 pre-trained chromatin states to the genome of FLS using chromHMM^13^. We identified Topologically Associating Domains (TADs) and determined significant chromatin interactions. Finally, we incorporated RNA-seq data from FLS. These analyses provided a comprehensive annotation of the epigenome and transcriptome of FLS (Fig. 1 a, b).

**Table 1.**
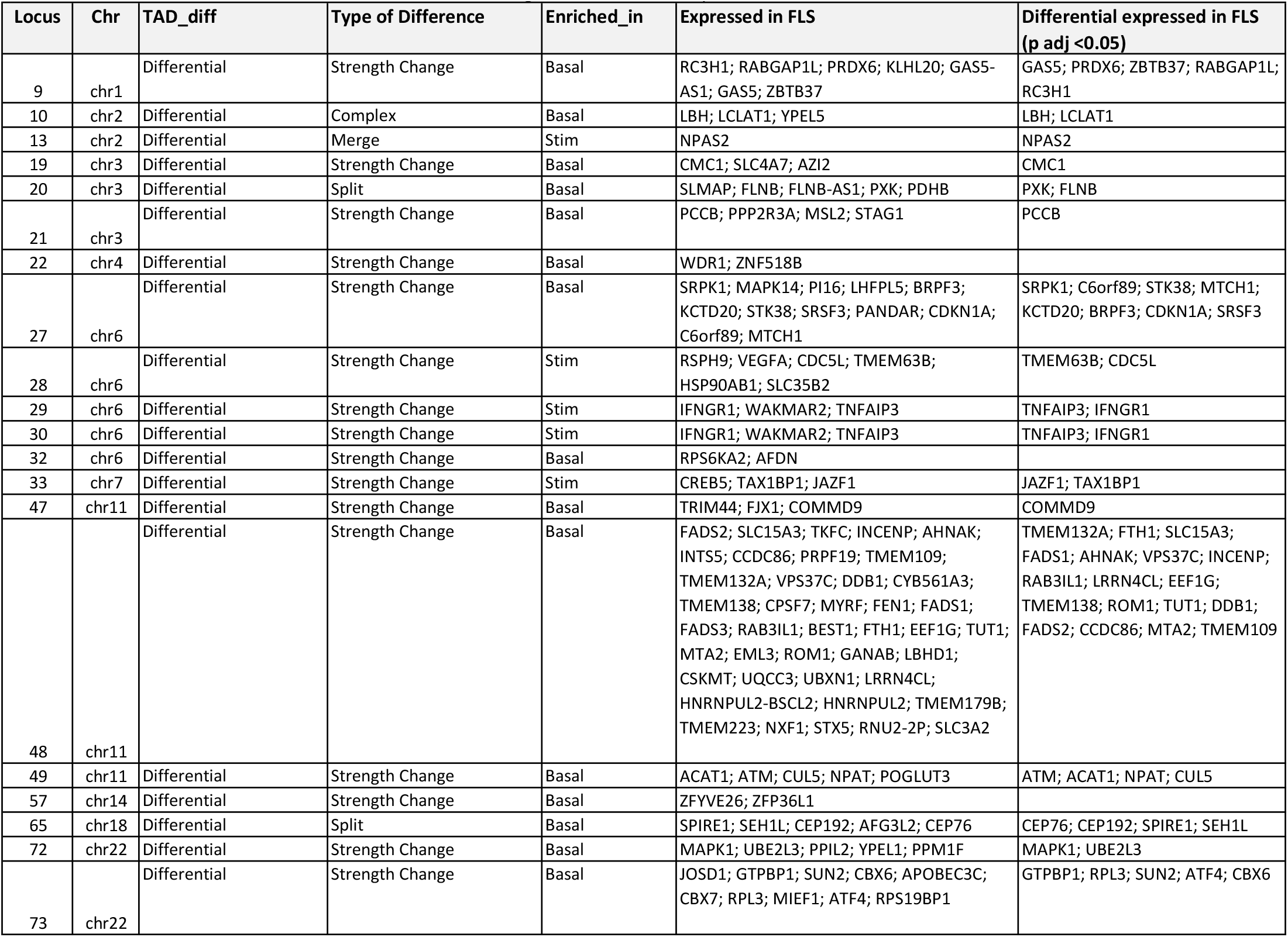
Alterations in TAD structure after TNF stimulation and genes in stimulation specific TADs

**Table 2.** Changes of gene expression in loci with changed chromatin interactions after TNF stimulation *Genes with a log2fold change >±0.5 and padj< 0.05 are marked in red (downregulated) and in green (upregulated)*

| Locus | Interacting Gene | Base Mean Expression | log2Fold Change | lfcSE | stat | p-value | padj |
| --- | --- | --- | --- | --- | --- | --- | --- |
| 12 | SPRED2 | 772.22 | -0.34 | 0.08 | -4.36 | 1.29E-05 | 7.23E-05 |
| 19 | SLC4A7 | 3702.62 | -0.23 | 0.13 | -1.80 | 0.072 | 0.145 |
| 20 | PXK | 1012.38 | -0.33 | 0.11 | -3.05 | 0.002 | 0.008 |
| 23 | RBPJ | 5319.00 | -0.56 | 0.22 | -2.51 | 0.012 | 0.032 |
| 24 | IL6ST | 18850.31 | 0.72 | 0.08 | 8.91 | 4.91E-19 | 2.30E-17 |
| 30 | TNFAIP3 | 7841.42 | 4.57 | 0.19 | 23.43 | 2.14E-121 | 3.51E-118 |
| 34 | CDK6 | 4906.40 | 0.92 | 0.12 | 7.89 | 3.05E-15 | 9.21E-14 |
| 35 | TNPO3 | 1671.71 | 0.05 | 0.07 | 0.76 | 0.449 | 0.597 |
| 41 | PHF19 | 1065.75 | 0.71 | 0.14 | 5.09 | 3.56E-07 | 2.74E-06 |
| 42 | TRAF1 | 1041.51 | 2.80 | 0.22 | 12.65 | 1.14E-36 | 1.98E-34 |
| 48 | VPS37C | 552.68 | 0.71 | 0.07 | 10.56 | 4.76E-26 | 3.90E-24 |
| 52 | CDK2 | 648.74 | 0.85 | 0.13 | 6.69 | 2.23E-11 | 3.72E-10 |
| 52 | RAB5B | 1805.92 | -0.24 | 0.06 | -4.06 | 4.88E-05 | 2.42E-04 |
| 52 | RNF41 | 1878.98 | -0.63 | 0.11 | -5.62 | 1.93E-08 | 1.88E-07 |
| 52 | ANKRD52 | 1954.38 | 0.23 | 0.07 | 3.09 | 0.002 | 0.007 |
| 52 | CNPY2 | 1033.14 | 0.37 | 0.07 | 5.30 | 1.16E-07 | 9.89E-07 |
| 52 | PAN2 | 312.46 | -0.23 | 0.08 | -2.80 | 0.005 | 0.015 |
| 52 | STAT2 | 4951.27 | 0.74 | 0.20 | 3.77 | 1.64E-04 | 7.21E-04 |
| 52 | TIMELESS | 728.81 | 1.11 | 0.14 | 8.10 | 5.66E-16 | 1.87E-14 |
| 52 | GLS2 | 18.80 | 0.34 | 0.26 | 1.31 | 0.191 | 0.317 |
| 53 | SLC26A10 | 10.97 | -0.29 | 0.39 | -0.75 | 0.455 | 0.603 |
| 53 | OS9 | 5461.71 | -0.12 | 0.07 | -1.83 | 0.068 | 0.138 |
| 53 | AGAP2 | 5.83 | 0.43 | 0.31 | 1.36 | 0.173 | 0.294 |
| 53 | TSPAN31 | 370.51 | -0.29 | 0.11 | -2.56 | 0.010 | 0.028 |
| 53 | CDK4 | 1630.73 | 0.06 | 0.07 | 0.88 | 0.378 | 0.529 |
| 53 | AVIL | 64.47 | -0.44 | 0.15 | -2.93 | 0.003 | 0.011 |
| 53 | CTDSP2 | 3707.75 | -0.47 | 0.06 | -7.79 | 6.45E-15 | 1.85E-13 |
| 54 | SH2B3 | 1493.15 | 0.52 | 0.10 | 5.29 | 1.25E-07 | 1.06E-06 |
| 54 | ATXN2 | 781.23 | -0.05 | 0.08 | -0.71 | 0.478 | 0.625 |
| 64 | ORMDL3 | 800.27 | 0.40 | 0.08 | 4.75 | 1.99E-06 | 1.32E-05 |
| 64 | PSMD3 | 1842.20 | 0.18 | 0.05 | 3.66 | 2.53E-04 | 0.001 |
| 65;68 | IFNAR2 | 644.38 | 2.81 | 0.11 | 26.14 | 1.14E-150 | 2.92E-147 |
| 68 | IFNAR1 | 2355.18 | 0.95 | 0.08 | 12.55 | 3.95E-36 | 6.53E-34 |
| 68 | IFNGR2 | 1248.98 | 1.10 | 0.08 | 12.90 | 4.71E-38 | 9.03E-36 |
| 68 | ITSN1 | 1138.10 | -0.24 | 0.11 | -2.10 | 0.036 | 0.080 |

lfcSE = standard error for log2 fold change, stat = *statistic* value for the null hypothesis, padj = p-value adjusted for multiple testing using Benjamini-Hochberg

**Fig 1:**
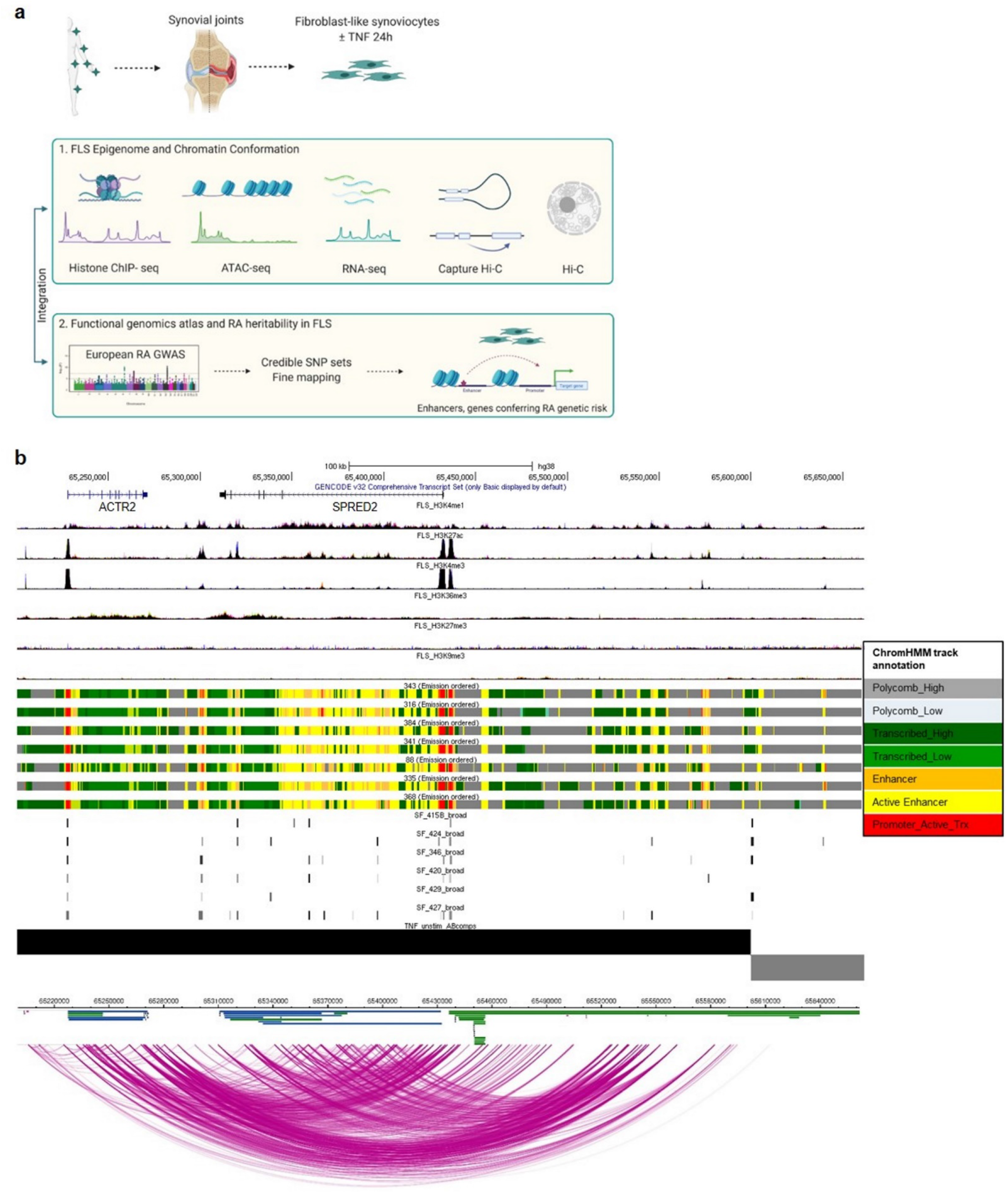
Epigenomic and 3D chromatin atlas of human FLS. **a)** Schematic representation of the workflow to comprehensively annotate the transcriptome, epigenome and chromatin structure of FLS and define their contribution to RA heritability. This figure was created using BioRender. **b)** The SPRED2 locus as an example genomic region demonstrating the annotation of epigenetic states and chromatin architecture in unstimulated FLS. Shown are from top to bottom, genes, ChIP-seq peaks (H3K4me1, H3K27ac, H3K4me3, H3K36me3, H3K27me3, H3K9me3), ChromHMM annotation in 7 different FLS lines, ATAC-seq peaks in 6 different FLS lines, A/B compartments (black bar open chromatin, grey bar closed chromatin), chromatin interactions.

We cross-validated the individual datasets to confirm the quality of the generated FLS data. The open chromatin regions as identified with ATAC-seq showed high enrichment of promoter chromatin states (Transcription Start Sites [TSS]) and active enhancers identified by chromHMM (Fig. 2a). In Capture Hi-C (CHiC), interactions of the selected prey fragments containing previously-reported lead SNPs at RA loci^2^ (details in Online Methods) were enriched for promoters (TSS), sites of transcription and enhancers (as defined by chromHMM) (Fig. 2b). At TAD boundaries, transcription and promoter states defined by chromHMM were enriched (Fig. 2c). Basal gene expression as measured by RNA-seq was highest in active TSS (Fig. 2d). Taken together, these analyses validated that we accurately captured chromatin states and chromatin interactions in FLS and that we have generated a comprehensive epigenetic and transcriptomic map of FLS genomes.

**Fig 2:**
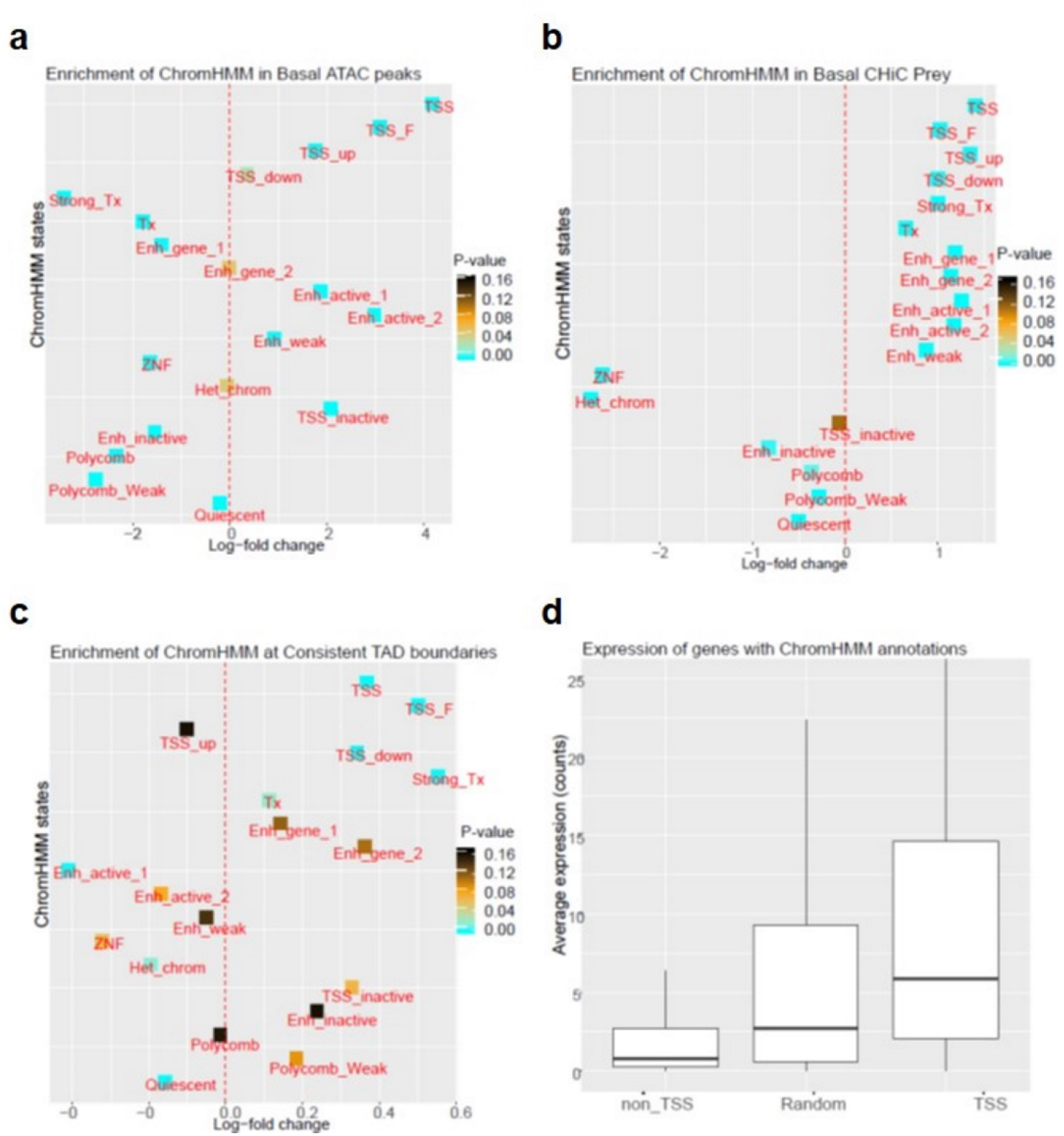
Cross validation of generated datasets defining the chromatin landscape of FLS. **a)** Log fold change enrichment of chromatin states as defined by ChromHMM in ATAC-seq data. **b)** Log fold change enrichment of chromatin states as defined by ChromHMM in prey fragments of Capture HiC measurements. **c)** Log fold change enrichment of chromatin states as defined by ChomHMM in consistent TAD boundaries. **d)** Basal average expression of genes (RNA-seq counts) across non-TSS, TSS and random ChromHMM annotations. TSS = transcription start site, TSS_F = flanking TSS; TSS_up = upstream TSS; TSS_down = downstream TSS; Enh_gene = enhancer genic; ZNF = zinc finger; Het_chrom = heterochromatin

### TNF induces changes in chromatin organization that correspond to altered gene expression in stimulated FLS

To explore the effect of a pro-inflammatory environment on the chromatin landscape of FLS, we performed Hi-C, CHiC, ATAC-seq and RNA-seq experiments in FLS with and without stimulation with TNF (Supplementary Table 2).

We first computed changes in A/B compartments, which are large, cell-type specific organisational units of the genome, associated with chromatin activity (A = open chromatin, B = closed chromatin)^14^. 94.8% of A and 95.7% of B compartments were consistent between basal and stimulated FLS. Small changes in A/B compartments after stimulation are expected, as A/B compartments infer chromatin activity at DNA segments in low resolution. One of the genomic regions that changed from an inactive (B) to an active (A) compartment on TNF stimulation contains RA associated variants that interact with the *TNFAIP3* gene.

We then explored the influence of TNF on the organization of TADs in FLS. Genes within the same TAD tend to be co-regulated and gene promoters and enhancers often interact within the same TAD^15^. Changes in TAD boundaries, as found in cancer, can induce major changes in gene expression^15^. We compared TADs between basal and stimulated FLS with TADCompare^16^. TADCompare classifies TADs as either non-differential or differential based on changes in position or strength of TAD boundaries. Between our conditions, we identified an average of 4,116 TAD boundaries in FLS samples. Whilst the vast majority (3259, 79.2%) of TAD boundaries were unchanged between basal and stimulatory conditions, 20.8% of differential TAD boundaries exhibited change in position (9.15 % boundary change complex, split, merge or shifted) or strength (11.66 % differential boundary magnitude) (Fig. 3a), indicating that the FLS genome exhibits changes in 3D structure upon TNF stimulation.

**Fig 3.**
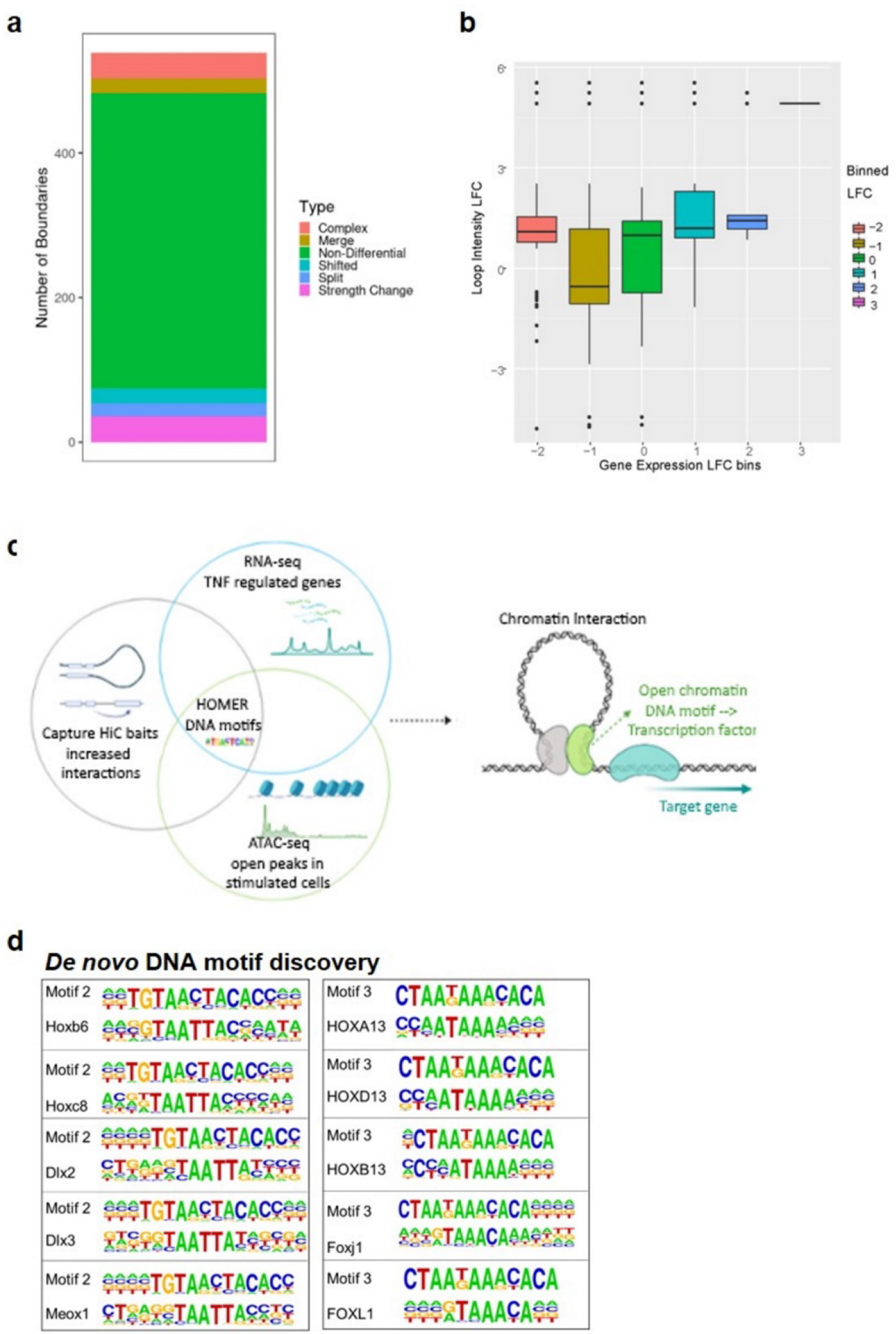
Effect of TNF stimulation on the chromatin landscape in FLS. **a)** Analysis of TADs boundaries after TNF stimulation in FLS by TADCompare. Number of non-differential (green) and differential TAD boundaries is shown. Differential TAD boundaries are classified as boundary position changes (complex, merge, shifted, split) or strength change (differential boundary magnitude). Complex, merged and split boundary changes represent the most disruptive changes of the 3D structure of the genome. **b)** Correlation of the loop intensity as determined by CHiC with change in the expression of nearby genes (log fold change). **c)** Graphical representation of the RNA-seq, ATAC-seq and CHiC data integration to identify transcription factor binding sites in TNF-stimulated FLS. This figure was created using BioRender. **d)** De novo DNA motif discovery identifies two motifs (motif 2 and motif 3) with high similarity to the binding sites of homeobox (TAATTA) and forkhead box transcription factors (TAAA) in the dataset with TNF-repressed genes.

By analysing CHiC data (details in Online Methods), we observed around 800 quantitatively differentially interacting regions between basal and stimulated FLS. The intensity of the differential interactions between the regions correlated with the fold-change of expression of the interacting genes (Fig. 3b). Notably, interaction strength increased after stimulation for genes with differential expression, irrespective of whether expression increased or decreased after stimulation, thereby suggesting that chromatin interactions influence activating and repressive TNF transcriptional responses in FLS (Fig. 3b).

To further explore the regulation of gene transcription after TNF stimulation, we focused on CHiC baits and prey that exhibited increased interaction strength with regulated genes after stimulation. We overlapped these regions with the measurements of open chromatin peaks (ATAC-seq) in stimulated cells. We then used Hypergeometric Optimization of Motif EnRichment (HOMER) to detect known transcription factor binding sites (TFBS) or DNA motifs with high similarity to known TFBS, that were overrepresented at the sites with open chromatin, increased chromatin interactions and differential gene expression compared to random background sequences (chosen by HOMER) (Fig. 3c).

Enrichment analysis of known TFBS in open chromatin identified TPA response elements (TREs; TGA(G/C)TCA) as the most enriched motif in the data sets with increased as well as decreased gene expression (Supplementary Dataset 2). TPA response elements serve as canonical binding sites for the subunits of the Activator Protein-1 (AP-1) transcription factor that is closely linked to the pathogenesis of RA^17^. The different subunits of the AP-1 family form homo- and heterodimeric transcription factor complexes with distinct activating and repressing functions^18^. Open chromatin sites with increased CHiC interactions, but decreased gene expression in stimulated FLS were additionally enriched for BACH2 (broad complex-tramtrack-bric a brac and Cap’n’collar homology 2) binding sites (Supplementary Dataset 2). Like AP-1, BACH2 belongs to the basic region leucine zipper (bZIP) family, but has a slightly different DNA sequence binding site (TGCTGAGTCA) and has a bric-a-brac-tramtrack-broad-complex (BTB) domain, which specifically interacts with co-repressors to repress transcription^19^. BACH2 is a highly conserved repressor with a central function in terminal differentiation, maturation and activity of B and T cells^20^. Intronic SNPs within the *BACH2* gene have been associated with the risk of different immune-mediated diseases, including RA^21,22^. Our data suggest that in addition to regulating immune cell functions, BACH2 may play a notable role in regulating the TNF response of FLS in RA.

De novo DNA motif discovery in the dataset with decreased levels of gene expression after TNF stimulation was enriched for two DNA motifs (DNA motifs 2 and 3) with high similarity to binding sites for several homeobox and forkhead box proteins (Fig. 3d). Together, they were present in 5.5% of the gene sites (motif 2: 3.7%, background 0.2%; motif 3: 1.8%, background 0%; p-value = 10^−12^). These results indicated a potential role for these developmental transcription factors in transcriptional repression of genes in TNF stimulated FLS. Since some of these transcription factors are exclusively expressed in FLS at distal joint locations (HOXA13, HOXD13)^8^, this suggests that TNF responses of FLS may be different at specific joint locations.

In summary, by combining CHiC, ATAC-seq and RNA-seq analyses we confirmed the activating and repressive actions of AP-1 in regulating the TNF response of FLS and we suggest that developmental transcription factors can serve as potential novel repressors of transcriptional response to TNF in FLS.

### FLS and immune cells are drivers of RA heritability

We then used the generated knowledge on regulatory DNA elements in FLS to quantify the heritability of RA that can be attributed to active regulatory DNA elements in FLS. We computed the partitioned heritability^23^ in FLS and other cell types (HLA regions excluded; details in Methods section). Epigenetic data for non-FLS cell types were acquired from published datasets^11^. We defined active regulatory elements of the genome as the union of H3K4me1, H3K4me3 and H3K27ac peaks, as these histone modifications are associated with transcriptional activity and enhancer/promoter elements. With this approach, we estimated that 12%-24% of the non-HLA RA heritability can be attributed to the active DNA regulatory elements in FLS samples (Fig. 4a).

**Fig 4.**
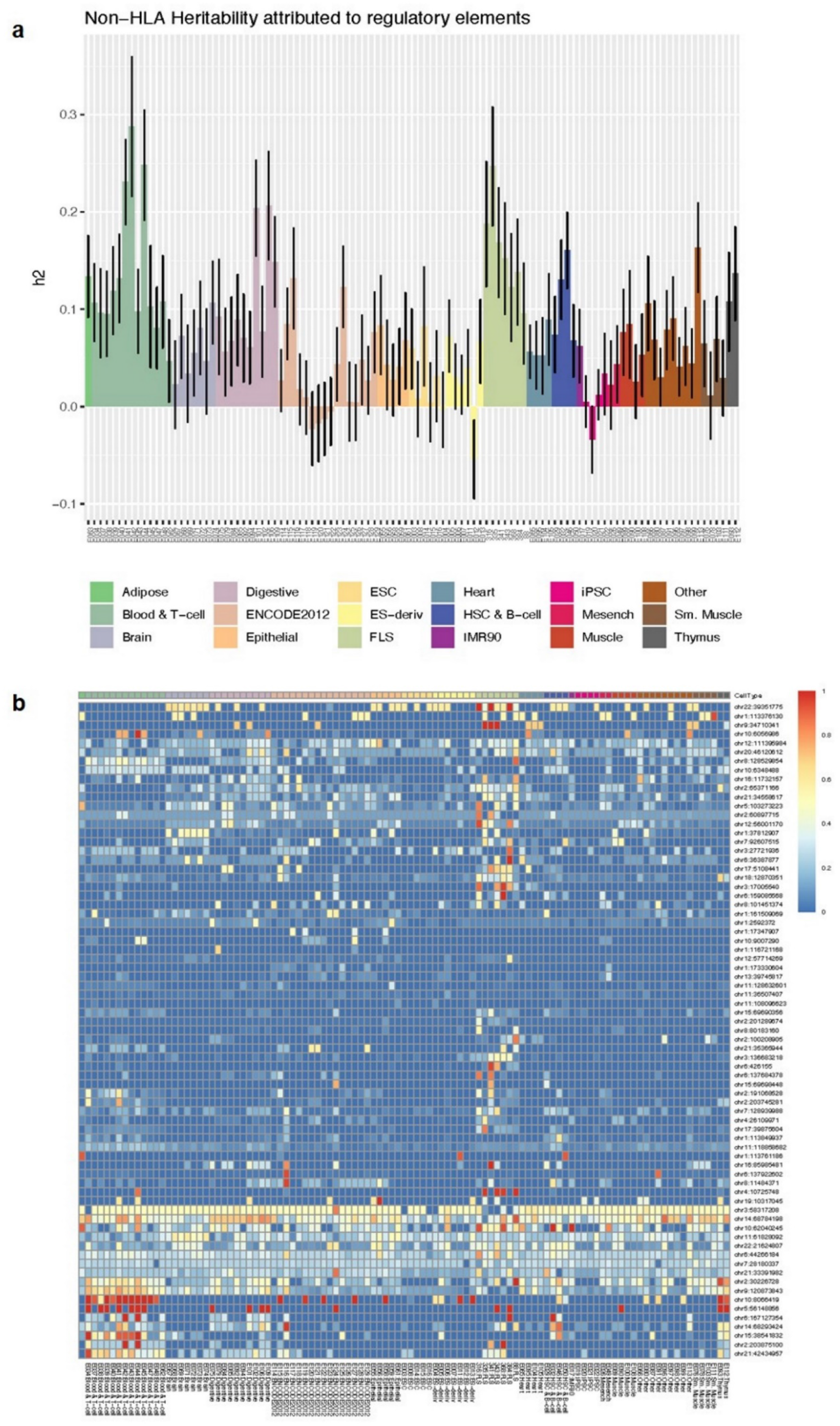
Heritability and causal SNPs in FLS. **a)** Partitioned heritability (h2) of RA attributed to active regions in each sample of FLS (n=7) and 111 available Roadmap cell types/tissues (Epigenomics Mapping Roadmap Consortium^11^). **b)** The sum of posterior probability overlapping active DNA regulatory elements at each of the 73 sites, defined as the union of H3K4me3, H3k4me1 and H3K27ac marks, across FLS samples and the 111 cell types/tissues.

We then considered RA risk loci attaining genome-wide significance (*p*<5×10^−8^) in the European ancestry component of the largest published trans-ethnic RA GWAS meta-analysis (HLA regions excluded)^2^. Where lead SNPs at genomic loci mapped within 1 Mb of each other, the loci were merged (Supplementary Table 3). Using approximate conditional analyses implemented in GCTA^24^, we identified 73 distinct signals of association with RA at locus-wide significance (*p*<10^−5^), with each signal being potentially driven by different underlying causal variants (Supplementary Table 4). For each signal, we performed fine-mapping to derive credible SNP sets that together account for ≥99% of the posterior probability of causality for the RA association. Across all 73 signals, the RA credible sets included a total of 8,787 variants, of which 2,654 variants had posterior probability of causality >0.01% (Supplementary Table 5). We then overlapped these 2,654 RA credible SNPs with the FLS epigenome and identified 274 SNPs mapping to active DNA regulatory elements in FLS (Fig. 4b). We also calculated the total posterior probability found within active DNA regulatory elements across the credible SNP sets for 111 primary cell types and tissues, whose epigenomes were published by the Roadmap Epigenomics Mapping Consortium^11^ (Fig 4b). As expected, several credible sets exhibited high posterior probability in active DNA regulatory elements from B and T cells (n=35), of which some (n=14) also overlapped active DNA regulatory regions in FLS (Fig 4b, Supplementary Table 5 column R). Intriguingly, we identified several credible sets that were active in FLS only, but not in B and T cells (n=9; Fig. 4b, Supplementary Table 5 column R). In summary, these analyses showed that both immune cells and FLS mediate the effects of association signal at risk loci and contribute notably to the heritability of RA.

**Table 3.** Posterior probablity and cellular context of category 1 and 2 loci

| Locus | Chr | Number of credible SNPs (>posterior prior 0.001) | Posterior probability top SNP | total posterior probability top 3 SNPs | SNP category | Number of SNPs in 30% of FLS enhancers (posterior>0.001) | Number of SNPs in 30% of T cell enhancers (posterior>0.001) | Number of SNPs in 30% of B cell enhancers (posterior>0.001) | FLS, T, B, ALL, NONE (based on posteriors of SNPs in enhancers) | Top posterior rs numbers |
| --- | --- | --- | --- | --- | --- | --- | --- | --- | --- | --- |
| 4 | chr1 | 10 | 0.53 | 0.94 | 1 | 1 | 0 | 0 | FLS | rs4839319, rs4839318, rs77227025 |
| 5 | chr1 | 2 | 0.91 | 0.99 | 1 | 0 | 0 | 0 | B CELL | rs6679677, rs2476601 |
| 7 | chr1 | 10 | 0.25 | 0.62 | 1 | 0 | 0 | 0 | NONE | rs624988, rs771587, rs12137270, rs12405671, rs11586238 |
| 10 | chr2 | 9 | 0.21 | 0.59 | 1 | 2 | 3 | 2 | ALL | rs10175798, rs10173253, rs906868, rs7579944, rs1355208 |
| 17 | chr2 | 4 | 0.43 | 0.92 | 1 | 0 | 2 | 0 | T CELL | rs231724, rs231723, rs231775 |
| 20 | chr3 | 13 | 0.34 | 0.82 | 1 | 4 | 2 | 3 | ALL | rs73081554, rs185407974, rs180977001, rs35677470, rs114584537 |
| 24 | chr5 | 1 | 1 | 1 | 1 | 0 | 1 | 0 | T CELL | rs7731626 |
| 29 | chr6 | 8 | 0.23 | 0.67 | 1 | 0 | 0 | 0 | NONE | rs17264332, rs11757201, rs6920220, rs6927172 |
| 30 | chr6 | 15 | 0.42 | 0.86 | 1 | 2 | 5 | 3 | NONE | rs58721818, rs61117627 |
| 32 | chr6 | 9 | 0.32 | 0.71 | 1 | 0 | 5 | 0 | T CELL, B CELL | rs1571878, rs3093017, rs10946216 |
| 40 | chr9 | 3 | 0.37 | 0.98 | 1 | 0 | 0 | 0 | NONE | rs11574914, rs2812378, rs10972201 |
| 42 | chr10 | 6 | 0.73 | 0.87 | 1 | 0 | 0 | 0 | T CELL | rs706778, rs10795791 |
| 44 | chr10 | 10 | 0.21 | 0.49 | 1 | 0 | 10 | 0 | T CELL | rs537544, rs568727, rs570613, rs570730, rs7897792 |
| 46 | chr10 | 10 | 0.37 | 0.73 | 1 | 9 | 1 | 0 | ALL | rs12764378, rs71508903, rs77509998 |
| 60 | chr15 | 5 | 0.26 | 0.74 | 1 | 0 | 0 | 0 | NONE | rs919053, rs8026898, rs7170107, rs16953656, rs8043362 |
| 62 | chr16 | 9 | 0.29 | 0.82 | 1 | 0 | 0 | 0 | B CELL | rs13330176, rs2139492, rs2139493 |
| 66 | chr19 | 2 | 0.6 | 1 | 1 | 0 | 0 | 0 | NONE | rs74956615, rs34536443 |
| 70 | chr21 | 19 | 0.56 | 0.93 | 1 | 3 | 1 | 0 | FLS | rs8133843, rs8129030, rs9979383 |
| 73 | chr22 | 9 | 0.29 | 0.74 | 1 | 0 | 0 | 2 | B CELL | rs2069235, rs909685, rs9611155 |
| 1 | chr1 | 81 | 0.29 | 0.38 | 2 | 1 | 1 | 1 | NONE | rs187786174, rs60733400, rs876938 |
| 2 | chr1 | 13 | 0.29 | 0.52 | 2 | 0 | 0 | 0 | NONE | rs2240336, rs12737739, rs13375202 |
| 3 | chr1 | 36 | 0.43 | 0.51 | 2 | 1 | 0 | 0 | FLS | rs28411352, rs28489009, rs2306627 |
| 6 | chr1 | 29 | 0.22 | 0.42 | 2 | 0 | 2 | 4 | B CELL | rs1217404, rs2476604, rs1217420 |
| 11 | chr2 | 13 | 0.18 | 0.4 | 2 | 1 | 1 | 1 | ALL | rs34695944, rs56095903, rs67574266, rs13031237, rs13031721 |
| 13 | chr2 | 17 | 0.27 | 0.63 | 2 | 0 | 0 | 1 | NONE | rs9653442, rs6712515, rs11676922, rs1160542, rs10865035 |
| 18 | chr3 | 63 | 0.27 | 0.55 | 2 | 0 | 0 | 0 | NONE | rs4452313, rs4416363, rs7617779 |
| 19 | chr3 | 11 | 0.21 | 0.48 | 2 | 1 | 1 | 0 | T CELL, B CELL | rs9310852, rs4680838, rs9880772, rs1353286 |
| 22 | chr4 | 49 | 0.2 | 0.29 | 2 | 0 | 0 | 0 | NONE | rs7660626, rs13142500, rs6831973 |
| 31 | chr6 | 13 | 0.17 | 0.49 | 2 | 0 | 2 | 2 | B CELL | rs2451258, rs2485363, rs654690, rs1994564, rs212389 |
| 33 | chr7 | 47 | 0.24 | 0.47 | 2 | 3 | 1 | 2 | ALL | rs186735625, rs57585717, rs2158624 |
| 36 | chr8 | 20 | 0.08 | 0.23 | 2 | 3 | 1 | 0 | FLS, B CELL | rs2736337 |
| 38 | chr8 | 11 | 0.15 | 0.39 | 2 | 1 | 0 | 0 | FLS | rs678347, rs507201, rs657425 |
| 43 | chr10 | 17 | 0.27 | 0.45 | 2 | 0 | 3 | 2 | T CELL, B CELL | rs947474, rs10796038, rs10796040 |
| 45 | chr10 | 44 | 0.36 | 0.54 | 2 | 0 | 2 | 0 | NONE | rs12413578, rs144536148, rs186856025 |
| 47 | chr11 | 59 | 0.47 | 0.53 | 2 | 4 | 2 | 5 | NONE | rs12574838, rs331463 |
| 51 | chr11 | 17 | 0.12 | 0.36 | 2 | 2 | 8 | 0 | NONE | rs4936059, rs11221402, rs7106876 |
| 54 | chr12 | 22 | 0.22 | 0.51 | 2 | 0 | 1 | 1 | T CELL, B CELL | rs10774624, rs3184504, rs7310615 |
| 56 | chr14 | 12 | 0.13 | 0.37 | 2 | 3 | 2 | 0 | ALL | rs1950897, rs911263, rs1885013, rs2104047, rs3784099 |
| 57 | chr14 | 21 | 0.22 | 0.45 | 2 | 7 | 10 | 6 | ALL | rs7146217, rs36045050, rs11158764 |
| 58 | chr15 | 17 | 0.25 | 0.48 | 2 | 1 | 12 | 2 | T CELL, B CELL | rs8032939, rs8043085, rs4924273 |
| 61 | chr16 | 18 | 0.08 | 0.23 | 2 | 0 | 0 | 0 | NONE | rs11075010, rs1579258, rs4584833 |
| 63 | chr17 | 39 | 0.49 | 0.67 | 2 | 0 | 0 | 6 | NONE | rs7224929, rs58483057, rs2071456 |
| 67 | chr20 | 11 | 0.32 | 0.59 | 2 | 1 | 1 | 1 | ALL | rs4239702, rs4810485, rs1883832 |
| 71 | chr21 | 37 | 0.32 | 0.6 | 2 | 0 | 26 | 0 | T CELL | rs1893592, rs225433, rs11203203 |
| 72 | chr22 | 103 | 0.24 | 0.26 | 2 | 9 | 20 | 22 | T CELL, B CELL | rs11089637, rs11089620, rs5754387 |

**Table 4.**
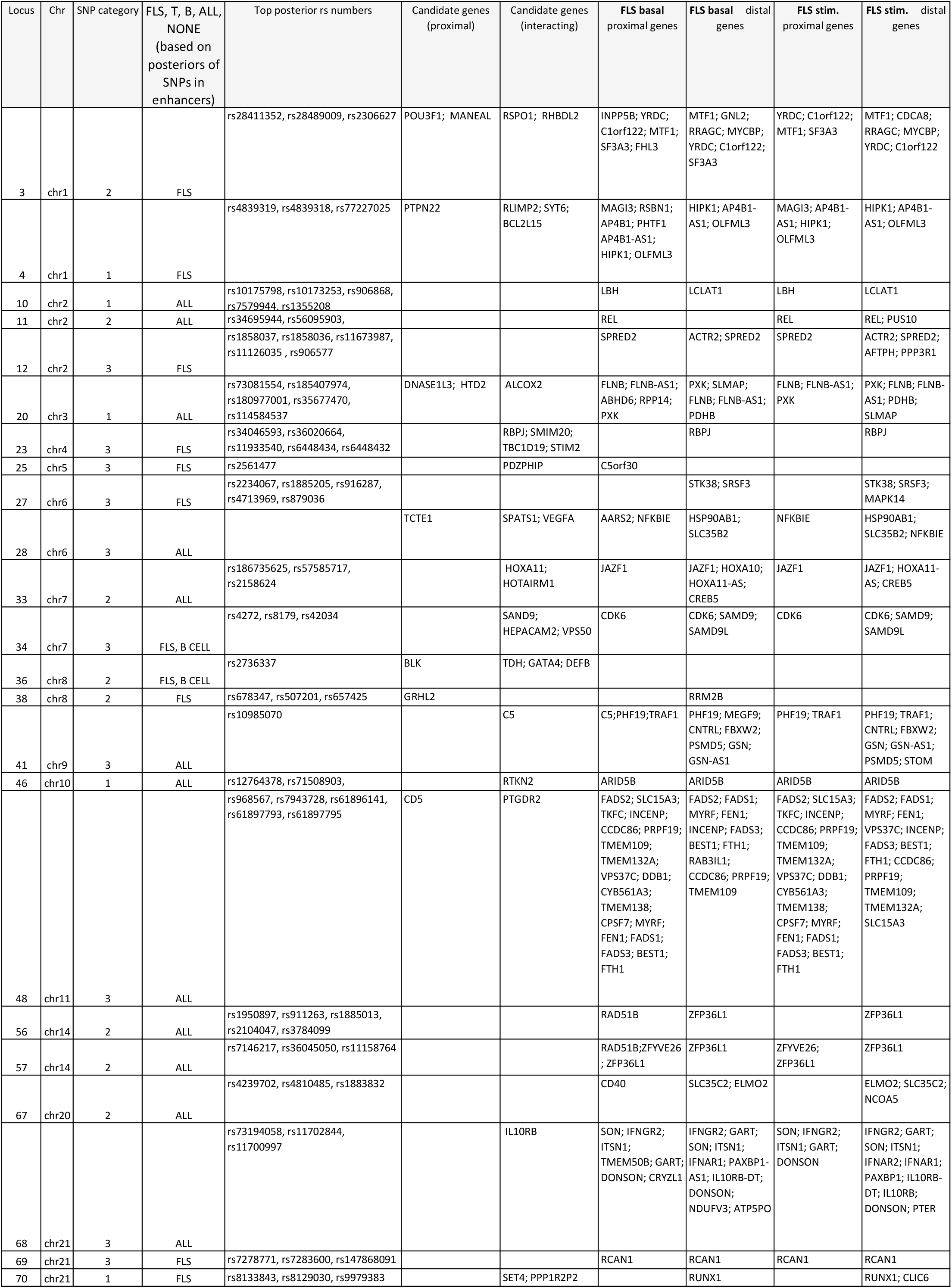
Loci which were assigned to FLS based on posterior probability

**Table 5.**
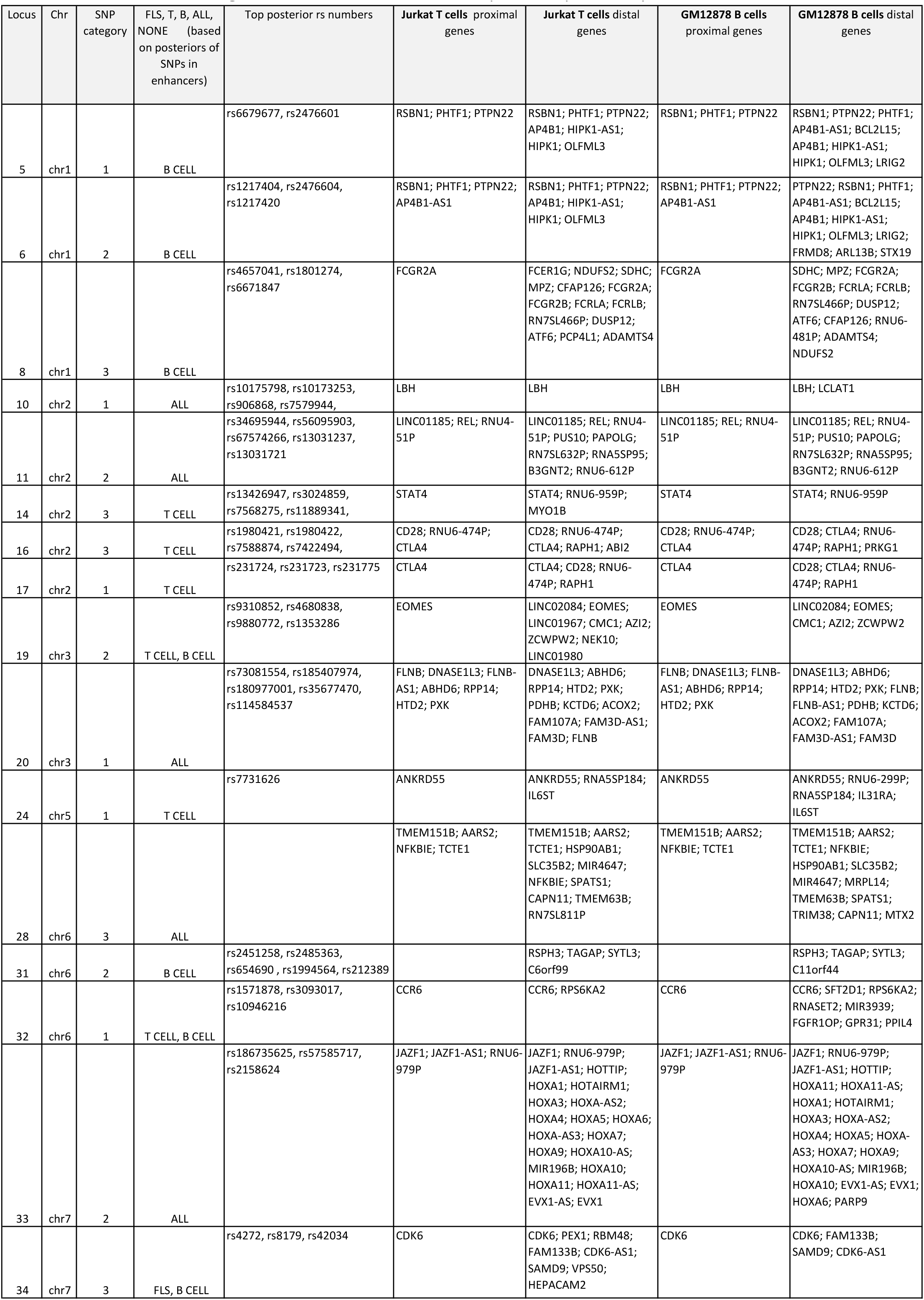
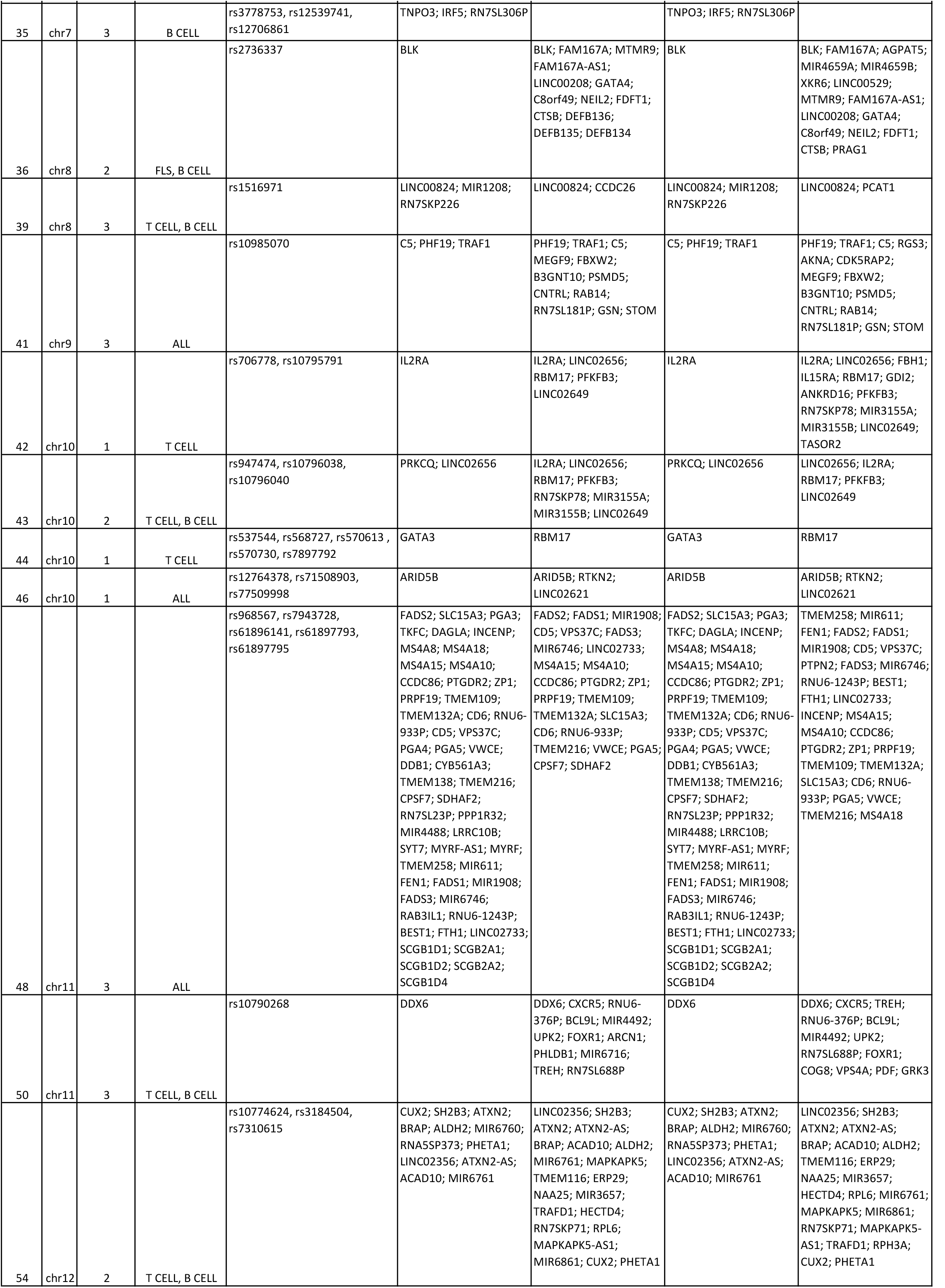
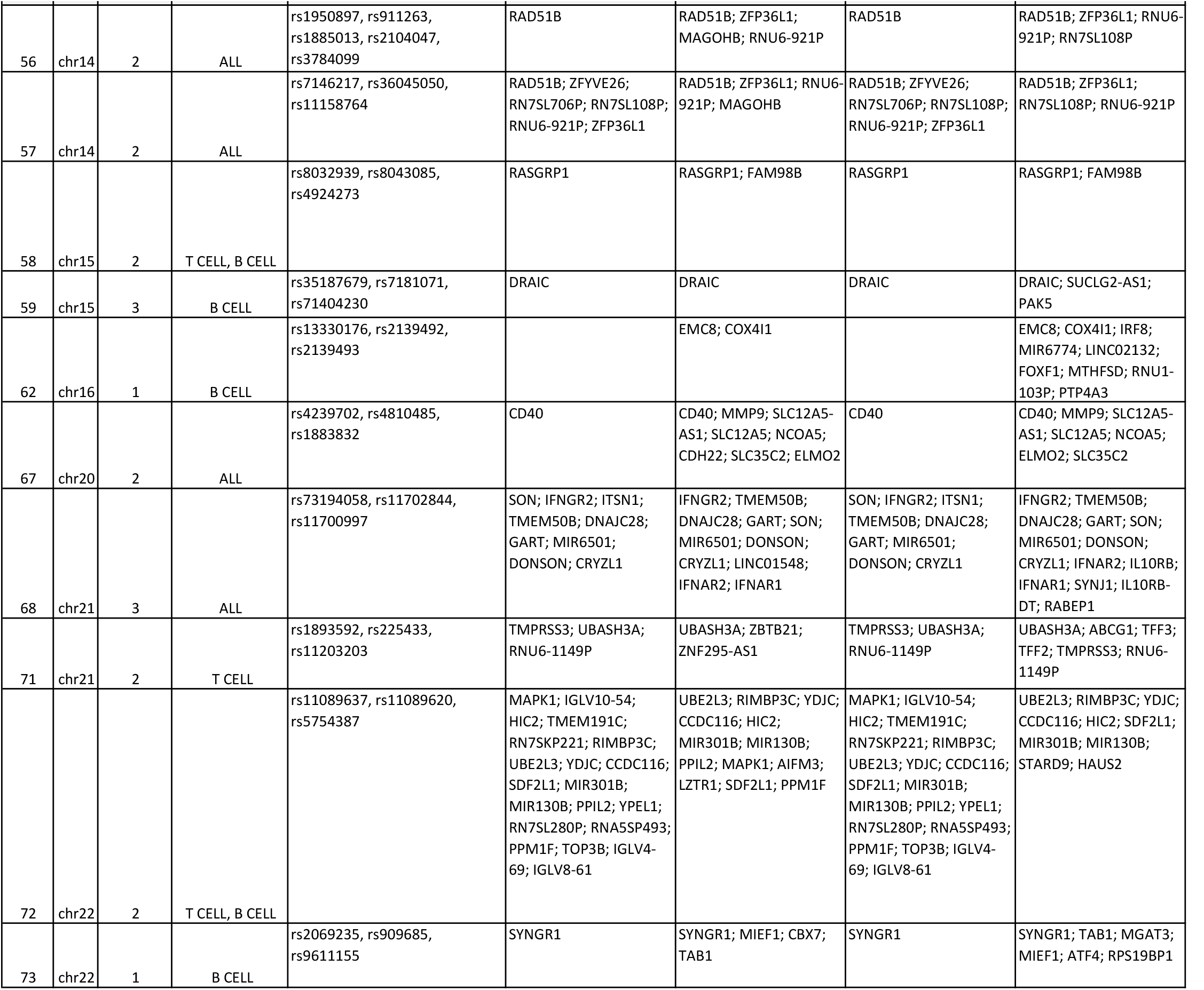
Loci which were assigned to T and/or B cells based on posterior probability

### TNF-induced alterations in 3D chromatin structure assign additional RA risk loci to FLS

To assign putative target genes in FLS at RA risk loci, we identified significant CHiC interactions between the region containing a credible SNP set (CHiC baits, see details in Methods) and a gene promoter. We defined gene promoters by downloading all transcripts from Ensembl (version 98) and assigning a 1000 base pair window directly upstream of each transcript as a promoter. In total, we determined 220,000 promoters for 57,602 genes, including non-coding RNA. Across RA risk loci, gene target assignments yielded a total of 228 and 227 interacting, expressed FLS target genes in basal and TNF stimulated conditions, respectively, with 188 gene targets shared between the conditions (Supplementary Table 5 columns W and X).

Since TADs have been shown to define probable limits of gene regulation, we overlapped TADs with RA credible sets. We observed that each credible SNP set is usually found within one or two adjacent TADs in unstimulated and stimulated cells. We then examined the genes within the TADs containing our credible SNPs in basal and stimulated FLS. The alterations in TAD boundaries between basal and stimulated FLS led to associated TADs overlapping different genes. Genes found within stimulation-specific TADs included *TNFAIP3, JAZF1, ZFP36L1, INFGR1* and *LBH*. Several of these genes, including *TNFAIP3, JAZF1, IFNGR1* and *LBH* also showed differential gene expression between basal and stimulated states (Table 1, Supplementary Table 5 columns AH and AI).

Furthermore, we observed a change in chromatin interactions as defined by CHiC in stimulated FLS at 17 RA risk loci, which were linked to 35 genes (Table 2). RNA-seq showed that the expression of 17 of the 35 genes was increased upon TNF stimulation in FLS (FDR < 0.05), 13 of which had a log2 fold change of > 0.5 (e.g. *TRAF1, TNFAIP3, IFNAR2)* (Table 2, Supplementary Figure 1). Nine of the 35 genes were downregulated after TNF in FLS (FDR < 0.05), two of them with a log2 fold change of > −0.5 (*RBPJ* and *RNF41*) (Table 2).

This correlated change in chromatin structure, interaction strength of RA implicated regions and gene expression upon stimulation, demonstrates how these loci are dynamic and active in FLS, and suggests that RA associated variants could thereby affect the transcriptional response to TNF in FLS.

### Epigenetic annotation of the fine-mapped SNPs in immune cells and FLS refines the putative causal credible set SNPs for more than 30% of the RA risk loci

Based on our genetic fine-mapping analysis, we identified three categories of RA risk loci. First, well characterised loci (“category 1”, n=19), where the credible set included ten or fewer SNPs or ≤ 3 SNPs contributing >80% of the posterior probability of causality. Second, loci with a localised signal (“category 2”, n=26), where the credible set included ≤ 20 SNPs with similar low posterior probabilities (0.1 - 0.2) or the lead credible SNP accounted for >20% of the posterior probability. Third, poorly characterised loci (“category 3”, n=28), where genetic fine mapping was largely ineffective, resulting in large (>20) credible SNP sets with equally negligible posterior probabilities (<0.05) (Supplementary Table 5 column K, Table 3). Examples of category loci 1-3 are shown in Supplementary Figures 2-4.

By mapping the credible SNP sets to the annotated active promoters and enhancers in T cells, B cells and FLS, we further refined nine of 19 category 1 loci to ≤ 3 credible SNPs in active enhancers in either immune cells (n=5 loci), FLS (n=1 loci) or both (n=3 loci) (Table 3, Supplementary Table 5 columns L, M, N). Similarly, we narrowed down the number of putative causal SNPs to ≤ 3 for 18 of the 26 category 2 loci, after mapping marks of chromatin activity to the credible set SNPs in immune cells (n=7 loci), FLS (n=3 loci) or both (n=8 loci) (Table 3, Supplementary Table 5 columns L, M, N).

Thus, by integrating genetic fine-mapping with functional chromatin annotation in immune cells and FLS, we identified 27 loci (37%) that harbour ≤3 putative causal RA risk variants having high posterior probabilities and mapping to cell type-specific active enhancers.

### Integrative analysis of genetic, expression and epigenetic datasets links putative causal genes and cell types

We then used our genetic fine-mapping and epigenetic datasets to determine the cell types in which credible SNPs at RA loci are active (FLS, immune cells, both, neither), the effector genes (proximal and interacting in FLS/immune cell types) and their expression in relevant cell types.

In total, 9 of the 73 loci were assigned exclusively to FLS, with 2 further loci assigned to FLS and B cells, and 12 to all three analysed cells types based on SNPs in cell-type specific enhancers (Table 4, Supplementary Table 5 column based on O, P, Q, labelled in column R). Credible SNPs in category 1 and 2 loci found predominantly in FLS active enhancers implicated genes including *GRHL2, MYCBP* and *RUNX1*. FLS assigned genes that were associated with category 3 risk loci, which showed negligible posterior probability (< 2%) in immune enhancer SNPs, included *SPRED2, RCAN1, CDK6* and *RBPJ* (Table 4). Notably, the 24 credible SNPs in the *RBPJ* locus and the 41 credible SNPs in the *CDK6* locus were reduced to just six and three SNPs, respectively, mapping to FLS specific enhancers. The *RBPJ* SNPs were localized in FLS specific enhancers, with none found in T or B cells (Supplementary Table 5, rs11933540). This indicated that the putative causal SNPs in the *RBPJ* locus might specifically affect the function of FLS in RA.

We then integrated the credible set SNPs with our previously established CHiC dataset from B cell (GM12878) and T cell (Jurkat) lines^12,25^. We found that the loci assigned to immune cell types associated with genes that are vital in T and B cell-specific activities (Table 5, Supplementary Table 5 columns AA to AD). Genes in category 1 and 2 loci, which associated with active immune cells enhancer regions included *CTLA4, IL2RA* and *GATA3* for T cells and *BLK* for B cells (Table 5, Supplementary Table 5 columns AA to AD). Of note, the *ANKRD55/IL6ST* locus (rs7731626 in Supplementary Table 5) had a single SNP in the credible list, an eQTL with both *ANKRD55* and *IL6ST*^26^ confined to an enhancer exclusive to T cells in our analysis. Immune cell assigned genes from category 3, where credible SNPs in immune enhancers accounted for >30% of the posterior probability, but had negligible posterior probability (< 5%) in FLS enhancers included *STAT4, CXCR5, CD28* and *MYC*.

These analyses highlighted a number of SNP-enhancer-gene combinations that could be assigned to an immune cell or fibroblast-driven risk of developing RA. We were able to assign >60% of the non-HLA RA loci with a putative causal cell type (FLS, B cells, T cells) and putative causal gene (column R not ‘none’). Compared to previous gene assignment results^2^, our method provides empirical evidence for an additional 104 RA associated genes at the 73 European risk loci.

### TNF stimulation induces major changes in chromatin organization of the TNFAIP3 and IFNGR2 risk loci with concomitant effects in the expression of interacting genes in FLS

Some of the risk loci emerged as particularly interesting in FLS, exemplifying how stimulation induced changes in chromatin conformation and gene expression can affect RA risk in FLS.

The intergenic region on chromosome 6q23 between *OLIG3* and *TNFAIP3*, which contains eight credible SNPs (rs17264332 in Supplementary Table 5), was dynamically linked to the *TNFAIP3* gene through DNA activity, chromatin interactions and gene expression. The organization of this genomic region changed from a closed, inactive (compartment B) to an open, active chromatin conformation (compartment A) upon TNF-stimulation of FLS (Fig. 5a), and TAD boundary strength increased in TNF-stimulated FLS (Table 3). These substantial alterations to the chromatin organization coincided with a strong increase in the expression of the interacting *TNFAIP3* gene in FLS (Fig. 5a, Table 2, Supplementary Figure 1).

**Fig 5.**
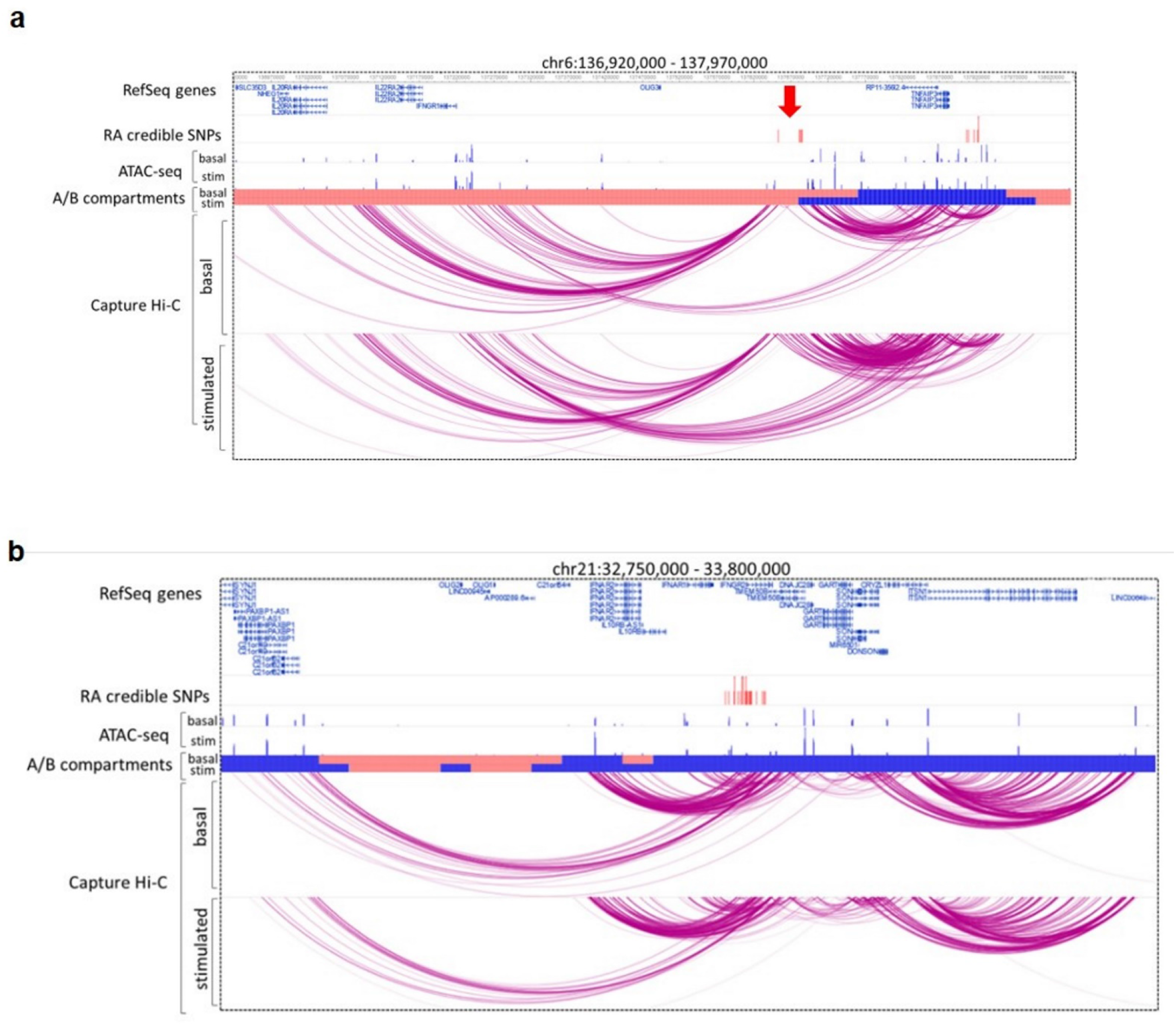
TNFAIP3 and IFNAR1 genetic risk loci - exemplary regions linked to TNF stimulation. Two exemplary risk regions where TNF stimulation had profound effects on chromatin structure and influenced the genetic regions containing RA SNPs. **a)** The TNFAIP3 region on chromosome 6q23 (red arrow) containing RA credible SNPs (red lines, rs17264332 in Supplementary Table 5) changed from closed chromatin (light red bar) to open chromatin state (blue bar) after TNF stimulation and exhibited increased interactions with the promoter of *TNFAIP3* in stimulated FLS. **b)** The genomic IFNGR2 region of the credible SNP set on chromosome 21 (rs73194058 in Supplementary Table 5) interacted with several nearby genes involved in the interferon response. These interactions were further enhanced by TNF stimulation. Chromatin at the IFNAR2 gene locus changed from a closed (light red bar) to open (blue bar) state in stimulated FLS.

Similarly, we demonstrated stimulation induced changes in chromatin activity in the *IFNGR2* region (rs73194058, Supplementary Table 5) in FLS. Our CHiC analysis showed that the credible set SNPs in this region interacted with several genes relevant to the interferon (IFN) pathway, such as *IFNAR2, IL10RB, IFNAR1* and *IFNGR2* (Supplementary Table 5 columns W to Z and Fig. 5b). TNF stimulation of FLS induced dynamic changes in chromatin interactions at this locus as assessed by CHiC and increased the expression of *IFNAR2, IFNAR1*, and *IFNGR2* (Supplementary Figure 1, Table 2, Fig. 5b). Additionally, chromatin activity in the region of *IFNAR2* changed (from inactive B to active A compartment) in stimulated FLS (Fig. 5b).

IFN pathways are strongly associated with the pathogenesis of RA and IFN-responsive genes are induced in FLS upon stimulation with TNF^27^. The *TNFAIP3/IFNGR1* region on chromosome 6 and the *IFNAR1/IFNGR2* region on chromosome 21 interacted with genes encoding five subunits of the IFN I/III receptors in FLS (Fig. 5 a,b), suggesting a close genetic link between FLS function and IFN response in RA.

### Genes linked to RA risk SNPs in FLS are functionally interlinked and regulate FLS-relevant RA functions

To predict biological processes influenced by potential transcriptional effects of risk variants active in FLS, we conducted analyses to predict protein-protein interaction, pathway enrichment and functional annotation clustering. For these analyses, we included all target genes of RA risk loci that were assigned to FLS as a causal cell type (“All” and/or “FLS” in column R of Supplementary Table 5).

We found significantly enriched protein-protein interactions for the genes in the loci active in FLS by using STRING protein-protein interaction networks (PPI enrichment p-value: <1.0e-016; Fig. 6a) and identified additional functional connections between the assigned genes by literature search. For instance, the transcription factor ZFP36L (rs1950897, Supplementary Table 5) negatively regulates the expression of CDK6^28^ by binding to the 3’UTR region of the *CDK6* gene, which contains the credible set SNPs at this locus (rs4272, Supplementary Table 5). CDK6 in turn interferes with DNA binding of Runx1^29^ (rs8133843, Supplementary Table 5). *ZFP36L, CDK6* and *RUNX1* were all assigned to FLS-active loci (Supplementary Table 5 column R), are functionally connected and regulate cell proliferation.

**Fig 6.**
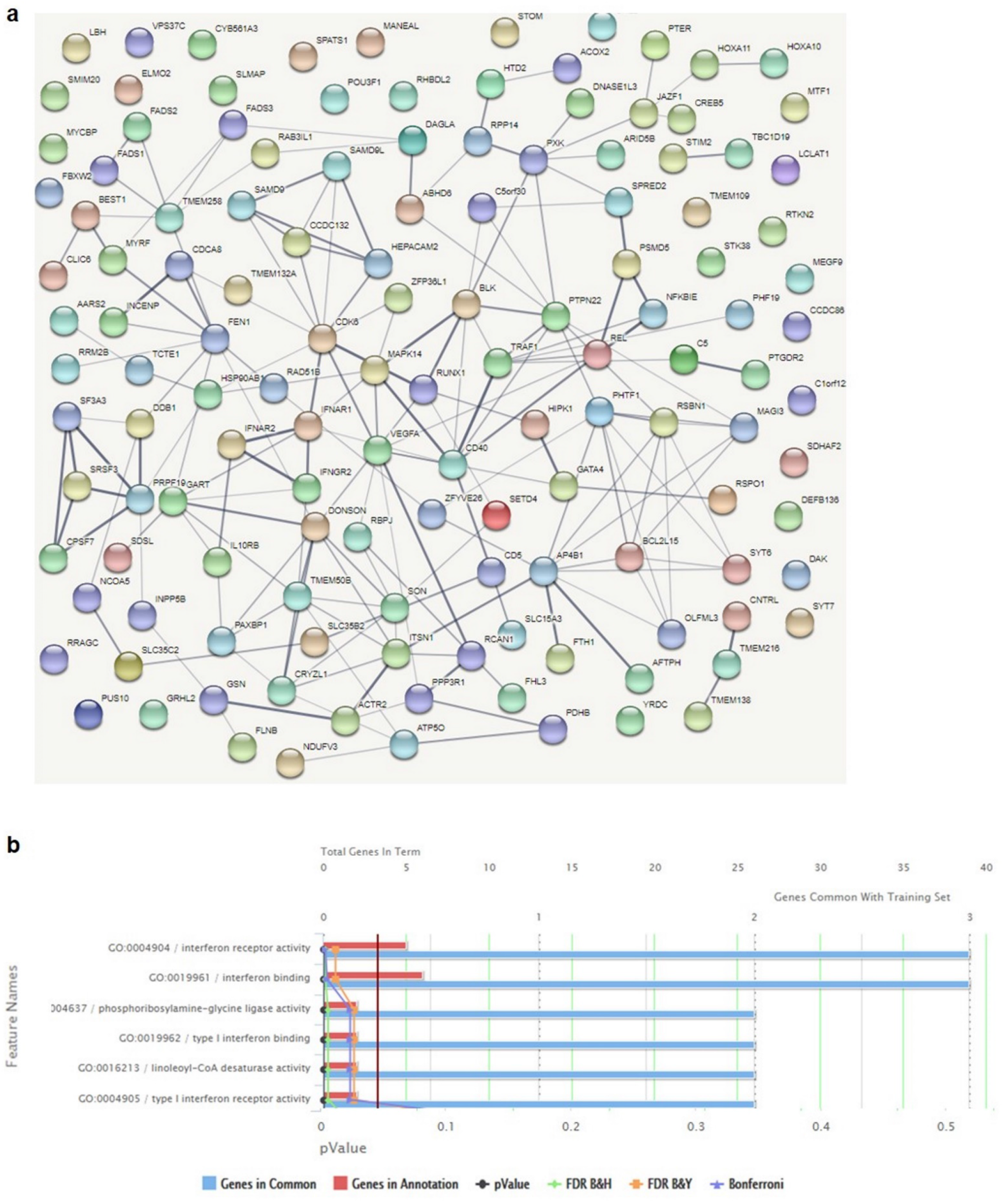
Predicted functional networks of genes that were associated with SNPs active in FLS. **a)** A protein-protein interaction network was established using STRING with default settings (medium confidence). The obtained network had more interactions than expected by chance with a protein-protein-interaction enrichment p-value of 1.28e-08. The thickness of the lines indicates the strength of data support. Colours, distances and location on the map were assigned randomly. **b)** Functional enrichment of genes interacting with SNPs active in FLS was detected using ToppFun in default settings. Significant terms for GO molecular function are shown. FDR = false discovery rate; B&H = Benjamini-Hochberg; B&Y = Benjamini-Yekutieli.

*CD40* (rs4239702 in Supplementary Table 5), *RBPJ* (rs11933540 in Supplementary Table 5) and *TRAF1* (rs10985070 in Supplementary Table 5) may constitute another genetically-influenced interlinked functional network in FLS. CD40 activation in FLS increased the expression of several cytokines relevant in RA, including VEGF and RANKL^30,31^. RBPJ (also known as CBF1), a regulatory transcription factor of the Notch signalling pathway, has been shown to repress the activation of CD40^32^. Similarly, TRAF1 can negatively regulate CD40 activity^33^.

Gene Ontology (GO) molecular function analysis (Fig. 6b) and functional annotation clustering of enriched pathways with the genes associated with credible set SNPs in FLS (Supplementary Table 5 column R and columns U-Z) revealed several clusters highly relevant to RA pathogenesis. These clusters included enrichment of genes involved in IFN response and viral defence (*IFNAR1, IFNAR2, CD40, IFNGR, C5, IL-10RB*) (Database for Annotation, Visualization and Integrated Discovery DAVID^34,35^ enrichment score 1.22) as well as lipid metabolism and fatty acid synthesis (*FADS1-3, ACOX2, LCLAT1, JAZF1, DAGLA*) (DAVID enrichment score 1.91). In addition, ‘cilium morphogenesis’ emerged as an enriched term (DAVID enrichment score 0.87) and several genes associated with RA risk SNPs in FLS were connected to the formation of the primary cilium (*C5orf30, GSN, TMEM138, TMEM216, CNTRL, INCENP, ACTR2*).

Overall, by integrating epigenetic and transcriptional data in FLS, we identified several functional relationships among RA risk variants and their target genes active in FLS. The multi-level effects of RA risk variants on key signalling pathways may contribute to the accumulated genetic risk in driving FLS activation and proliferation in RA.

### RA risk SNPs in the *RBPJ* enhancer region confer joint-specific genetic effects in FLS

Our epigenetic and functional analyses of the *RBPJ* locus identified *RBPJ* as a candidate causal and functional gene in FLS (rs11933540 in Supplementary Table 5). Mapping of the 24 credible SNPs to FLS enhancers in the *RBPJ* locus reduced the number of likely causal SNPs to six and rs874040 was identified as a strong candidate SNP for causality (Supplementary Table 5 rs11933540, Fig. 7a). To functionally establish that the rs874040-containing enhancer region can regulate the expression of *RBPJ*, we transduced FLS with lentiviral particles containing dCas9-VPR and two guide RNAs (g9 or g12) targeting the rs874040-containing enhancer region (Fig. 7a). FLS transduced with the activating dCas9-VPR and guide RNAs increased the expression of *RBPJ* compared to FLS transduced with the respective guide RNAs without dCas9-VPR (Fig. 7b). Even though the upregulation of *RBPJ* expression was modest (30%), which could be due to enhancer redundancy in this region, this experiment verified the regulation of *RBPJ* expression by the rs874040-containing enhancer region.

**Fig 7.**
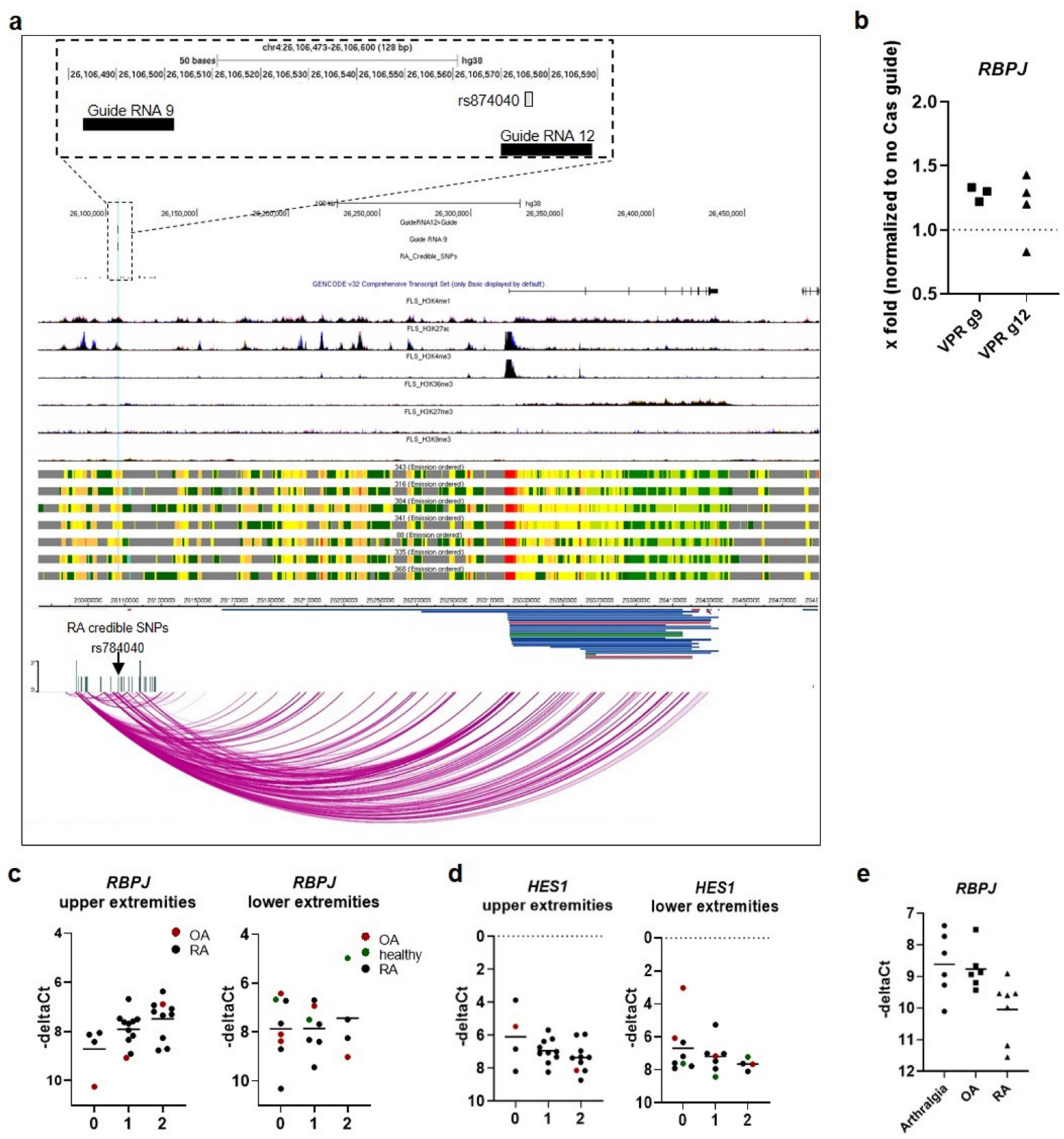
*RBPJ* expression in FLS is affected by genotype and disease. **a)** Fine mapping, epigenetic and chromatin conformation analyses at the *RBPJ* locus indicate rs874040 as a likely causal credible SNP and *RBPJ* as a candidate causal gene in this locus. The location of credible set SNPs with rs874040 and the guide RNAs (guide RNA 9 and guide RNA 12) targeting the rs874040-containing enhancer are indicated. **b)** *RBPJ* expression in FLS transduced with VP64-p65-Rta dCas9 (VPR) and two different guide RNAs (g9 and g12) targeting the genomic region around chr4:26106575 (rs87040). *RBPJ* expression was normalized to FLS that were transduced with respective guide RNAs but not VPR-dCas9 (set to 1). –deltaCt = cycle of threshold of *RBPJ* expression – cycle of threshold *RPLP0*. **c)** *RBPJ* expression in FLS isolated from individuals homozygous for rs874040 in the locus near the *RBPJ* gene (0), heterozygous (1) or homozygous for the wild type variant (2). Upper extremity joints included joints of the hand, elbows and shoulders; lower extremity joints included hips, knees and joints of the feet. **d)** Expression of *HES1* in the same FLS cohort. **e)** *RBPJ* expression in individuals with joint pain, but no histological signs of arthritis (arthralgia), OA and RA.

**Fig 8.**
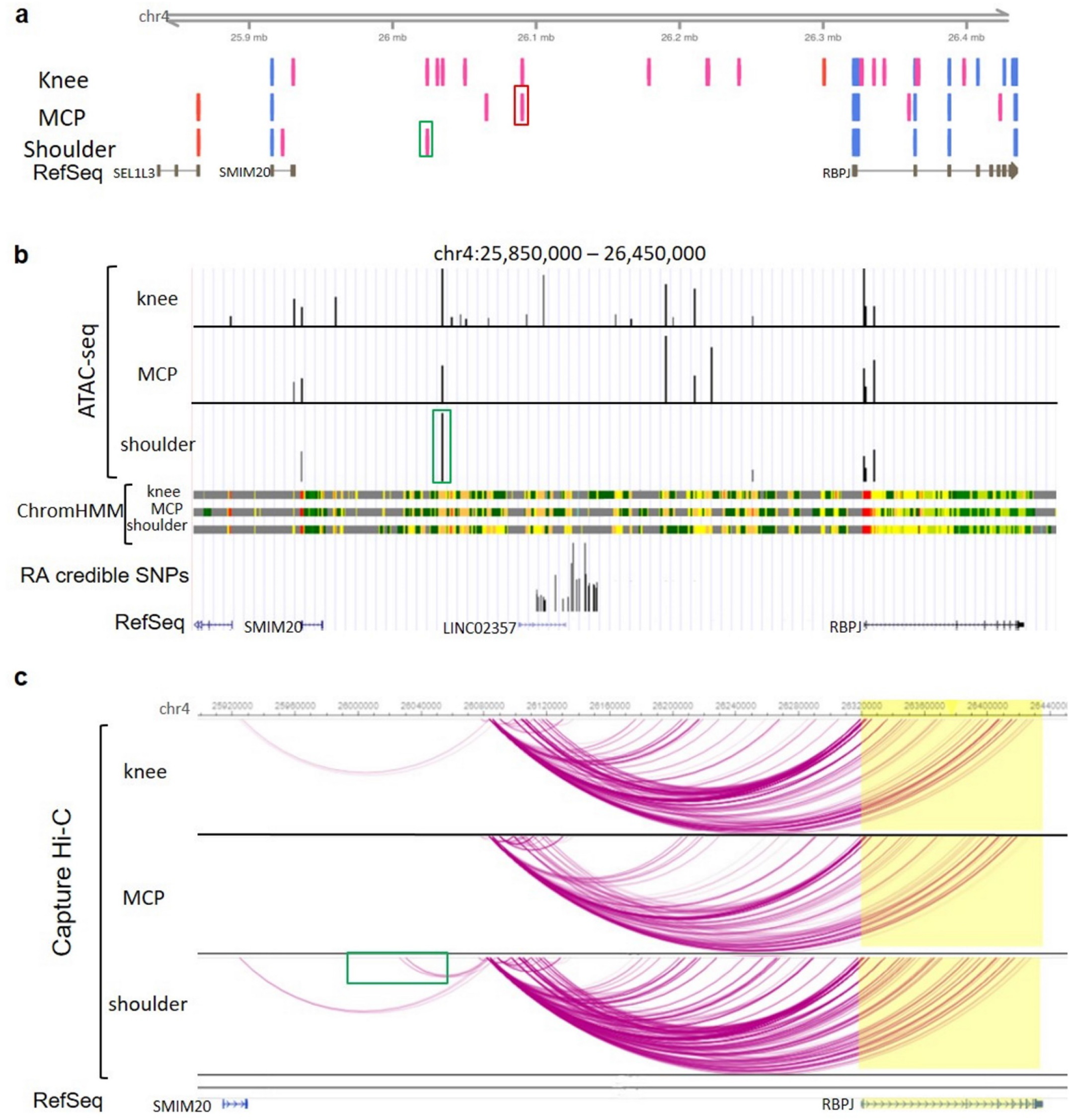
Joint-specific enhancers and chromatin interactions in the *RBPJ* locus might influence the joint-specific expression of *RBPJ*. **a)** CAGE measurements of active enhancers (pink bars) and active promoters (light blue bars) in FLS from knees (n=2), metacarpophalangeal (MCP) joints (n=3) and shoulders (n=2). Red box highlights enhancer used in MCP joints overlapping risk SNPs. Green box highlights main enhancer in shoulders. **b)** Representative depiction of open chromatin as measured by ATAC-seq (black bars) in FLS from knees (n=3), MCP joints (n=2) and shoulders (n=2) and representative depiction of ChromHMM regulatory regions in FLS from knees (n=3), MCP joints (n=2) and shoulders (n=2). Green box highlights main enhancer in shoulders. **c)** Chromatin interactions in FLS from knees (n=2), MCP joints (n=2) and shoulders (n=2) as measured by CHiC. Green box highlights interaction of the shoulder enhancer with the risk locus.

FLS homozygous for the risk allele of rs874040 exhibited lower expression of *RBPJ* mRNA compared to FLS with the wild-type variant. This effect was, however, present only in FLS from upper extremity joints, and not from lower extremity joints (Fig. 7c). It is known that RBPJ binds to the promoter of HES1 and represses its transcription^36^. Accordingly, the expression of *HES1* was increased in FLS from patients homozygous for rs874040 in upper extremity joints (Fig. 7d). TNF stimulation significantly downregulated *RBPJ* mRNA expression in FLS (Table 2), and FLS from RA patients expressed less *RBPJ* than FLS from patients with arthralgia (Fig. 7e). These data indicate that genetic predisposition and a pro-inflammatory environment can affect *RBPJ* expression in FLS, which might lead to increased activation of the Notch signalling pathway via HES1.

To explain the joint-specific effect of rs874040, we explored the enhancer landscape and the chromatin interactions in different upper and lower extremity joints. CAGE-seq data showed that the enhancer activity within the *RBPJ* locus is higher in knee FLS compared to shoulder and hand FLS (Fig. 7e). CAGE-seq enhancer signals largely overlapped with ATAC-seq peaks, being more abundant in knee FLS than in shoulder or hand FLS (Fig. 7f). Shoulder FLS appeared to mainly use an upstream enhancer that interacted with the *RBPJ* risk locus (green boxes, Fig. 7e). Overlap of ATAC-seq and CAGE-seq analyses was weaker in hand FLS, but CAGE-seq data indicated that hand FLS used an enhancer within the risk locus (red box, Fig. 7e). Additionally, chromatin interactions within the locus were generally weaker in hand FLS (Fig. 7g). Knee FLS activated several enhancers (Figs. 7e, f) and exhibited strong chromatin interactions across the locus (Fig. 7g). We analysed DNA-binding motifs in the enhancer overlapping the RA risk locus spanning chr4:26090045-26090465 (hg19) (red box in Fig. 7e) by using the JASPAR2020 database^37^. This enhancer contained TFBS for different HOX transcription factors (HOXA6, HOXA7, HOXA10, HOXB2, HOXB6, HOXB7, HOXB13, HOXD3, HOXD9, HOXD13), similar to the DNA motifs identified in open chromatin at repressed genes after TNF stimulation in FLS (Fig. 3d) and expressed in a joint-specific manner in FLS^38^.

Together these data suggest that joint-specific differences in chromatin interactions and enhancer usage could underlie the joint-specific effects of rs874040 on *RBPJ* expression in upper extremity joints. This illustrates that RA genetic risk can be different between the joints, thereby shaping a specific pattern of joint involvement in RA.

## Discussion

Deciphering the role of causal genetic variants underlying GWAS loci in RA, albeit challenging, provides an unbiased strategy to understand the core disease pathways and guide drug discovery^2,39^. Here we demonstrate that a significant proportion of the 73 European ancestry non-HLA RA risk loci contain disease-associated variants that are located within active regulatory DNA elements in FLS. Linking these DNA regions with target genes indicates genes and biological pathways that trigger RA susceptibility by stromal cell activation in the joint. Thus, we provide for the first time substantial evidence for an independent, causal role of FLS in RA genetic susceptibility and pathogenesis.

With our approach, we were able to assign RA risk loci to immune and/or stromal cells. Genes implicated in T cells, but not in FLS, showed a pattern of involvement in ‘canonical’ T-cell immunity, including *CTLA4, CD28, IL2RA*, and *GATA3*. Similarly, genes enriched in B cell specific enhancers were involved in B cell biology, including *IRF8, BLK*, and *TAB1*. The stromal activation observed in RA joints was clearly reflected in the predicted function of the identified FLS-specific regulatory variants, many of which were previously associated with RA pathogenesis. For example, several genes linked to RA credible SNPs in FLS were implicated in cell proliferation and tumour development (e.g. *SPRED2*^40^, *GRHL2*^41^, *CDK6*^*42*^, and *ZFP36L*^43^).

Most notably, the risk loci on chromosomes 6 (*TNFAIP3/IFNGR1*) and 2 (*IFNAR/IFNGR2*) could critically impact the contribution of FLS to the development of RA. Chromatin interactions in these regions connected RA risk variants with several genes encoding the subunits of type I (*IFNAR1, IFNAR2*), type II (*IFNGR1, IFNGR2*) and type III (*IL-10RB*) interferon receptors in FLS. Furthermore, they tightly linked the IFN response to TNF stimulation by interconnection with the *TNFAIP3* gene, encoding the TNF signalling repressor A20, and by their strong reaction to TNF stimulation. All three types of interferons signal via the JAK-STAT signalling pathway, which, along with TNF, is one of the central therapeutic targets in RA. A type I interferon gene signature is detectable in up to two thirds of patients with RA^44^ and it associates with an increased risk of developing RA as well as with therapeutic response to biological DMARDs like TNF inhibitors^45,46^. In FLS, TNF induces an extensive interferon gene response via secondary autocrine production of IFNβ and the activation of the IRF1-IFNβ-IFNAR-JAK-STAT1 axis^47,48^. Down syndrome (trisomy 21) leads to increased dosage of the IFN receptors encoded on chromosome 21, which results in a type I interferon gene signature with constant activation of interferon pathways in fibroblasts^49^. Notably, people with Down syndrome are at increased risk of developing erosive, inflammatory seronegative arthritis of their hands and wrists^50^. Together this strongly suggests a causal role for stromal activation of IFN pathways in the development of RA.

We showed that RA risk allele rs874040 is associated with reduced expression of *RBPJ* in FLS in a location specific manner. RBPJ, also called CBF1 or CSL, is a key transcriptional regulator of the Notch signalling pathway^51^. In the absence of Notch signalling, RBPJ represses Notch target genes (e.g. HES1). Upon activation of Notch signalling, RBPJ binds to the intracellular domain of the Notch receptor and enhances Notch-dependent gene expression. Loss of RBPJ leads to activation of dermal fibroblasts and promotes their transformation into cancer associated fibroblasts (CAFs), which play a crucial role in tumour development and growth^52,53^. Activation of Notch signalling was shown in RA FLS and induced FLS proliferation^54^. Furthermore, Notch signalling is critical for shaping the synovial environment by guiding the development of THY1+ sublining FLS, a subset of FLS that is expanded in RA synovial tissues^55^. Constitutive lower levels of RBPJ in FLS from individuals carrying the RBPJ risk variant could favour synovial enrichment of THY1+ sublining FLS, which are considered critical for the development of RA. Joint-specific differences in the chromatin landscape in this locus exemplify how genetic risk could result in the specific patterns of joint involvement that typically occur in chronic inflammatory joint diseases. Additionally, joint-specific expression of HOX transcription factors^8^, for which we suggest a role in gene repression after TNF stimulation in FLS, could contribute to joint-specific differences in the susceptibility to RA.

Further pathways that we found enriched in genes targeted by RA credible SNPs were connected to lipid metabolism and the primary cilium. FADS1 and 2 have been implicated in the production of anti-inflammatory unsaturated fatty acids in LPS-treated macrophages, contributing to the resolution phase of LPS-driven inflammatory response in macrophages^56^. Changes in the lipid metabolism have been suggested in RA FLS^57^, but specific functional data does not exist so far. The primary cilium serves as a hub for several cell signalling pathways, e.g. Notch^58^ and wnt signalling^59^. In FLS, it was shown that TNFR1 and TNFR2 localise to the cilium-pit^60^. The cilium connected proteins C5orf30 and GSN, that we found interacting with RA risk variants in FLS, were previously shown to be negative regulators of arthritis in mice^61,62^. Another ciliary protein, SPAG16 was found to be a genetic risk factor for joint damage progression in RA patients, increasing the production of matrix-metalloproteinases in FLS^63^. Future studies are required to demonstrate how changes in lipid metabolism and primary cilium affect the function of FLS and influence RA pathogenesis.

Overall, our research significantly advances the knowledge about putative causal SNPs, enhancers, genes and cell types affected by genetic risk loci in RA. Our analysis can direct future studies to investigate pathways that are genetically affected in a cell type specific way. This will ultimately enable the connection of an individual’s genetic risk with the causal pathways and cell types that drive disease, paving the way to stratified treatment decisions and precision medicine.

## Data Availability

The data is available from the authors upon request.

## Acknowledgements

This work was supported by the Wellcome Trust (award reference 207491/Z/17/Z), Versus Arthritis (award references 21754 and 21348) and by the NIHR Manchester Biomedical Research Centre. The views expressed are those of the author(s) and not necessarily those of the NHS, the NIHR or the Department of Health. CO and MFB were supported by the Swiss National Science Foundation (project 320030_176061) and the Georg and Bertha Schwyzer-Winiker Foundation.

## Contributions

X.G., M.F.B., A.P.M., P.M., S.E. and C.O. conceived and designed the study. M.F.B., K.K., A.M., T.K., B.B., G.O. and C.O. performed the experiments. X.G., M.H., R.M., A.P.M., P.M. and S.E. conducted fine mapping and bioinformatics analyses. M.M., A.F., C.D.B and O.D. collected patient samples and clinical data. X.G., M.F.B., A.P.M., P.M., S.E. and C.O. interpreted the data. X.G., M.F.B., A.P.M., S.E. and C.O. wrote the manuscript. All the authors critically discussed the manuscript and approved the submitted version.

## Online Methods

### Patients and cell culture

Synovial tissues were obtained from OA patients (n=9, 2male/ 8 female, mean age 68, range 54-88) and RA patients (n=30, 8 male/22 female, mean age 66, range 44-78) undergoing joint replacement surgery at the Schulthess Clinic Zurich, Switzerland. Patient’s characteristics are described in Supplementary Table 1. RA patients fulfilled the 2010 ACR/EULAR (American College of Rheumatology/European League Against Rheumatism) criteria for the classification of RA^1^. Samples from patients with joint pain without inflammation or cartilage destruction (healthy, 3 male/3 female, mean age 39, range 23-49) were obtained from the Queen Elizabeth Hospital in Birmingham, UK. The studies were approved by the local ethic committees of the University Hospital Zurich, Switzerland and the University of Birmingham, UK. Informed consent was obtained from all patients. Synovial tissues were digested with dispase (37 ° C, 1 h) and FLS were cultured in Dulbecco’s modified Eagle’s medium (DMEM; Life Technologies) supplemented with 10% fetal calf serum (FCS), 50 U ml−1 penicillin/streptomycin, 2 mM L-glutamine, 10 mM HEPES and 0.2% amphotericin B (all from Life Technologies). Purity of FLS cultures was confirmed by flow cytometry showing the presence of the fibroblast surface marker CD90 (Thy-1) and the absence of leukocytes (CD45), macrophages (CD14; CD68), T lymphocytes (CD3), B lymphocytes (CD19) and endothelial cells (CD31). Cell cultures were negative for mycoplasma contamination as assessed by MycoAlert mycoplasma detection kit (Lonza). FLS were used between passages 4-8. Information on the assays performed on each sample is given in Supplementary Table 2.

### RNA sequencing

RNA sequencing data from unstimulated samples (Supplementary Table 2) was retrieved from the European Nucleotide Archive (ENA) with the primary accession code PRJEB14422. A detailed description of sample preparation and sequencing procedures is given in Frank-Bertoncelj et al.^2^. For RNA sequencing of TNF stimulated FLS, cultured FLS were treated with 10 ng/ml human recombinant TNF (Roche) for 24 h or were left untreated. Total RNA was isolated using the RNeasy Mini kit (Qiagen) including on-column DNAase I digestion. Part of the libraries (n=12) were prepared using the NEB Next Ultra Directional RNAseq protocol with ribosomal depletion and were sequenced using Illumina HiSeq4000 with 75bp paired end reads. The additional libraries (n=20) were generated using the Illumina TruSeq Stranded total RNA protocol with the TruSeq Stranded total RNA Sample Preparation Kit and were sequenced using Illumina Novaseq 6000. All Fastq-files were mapped to hg19 and sequence reads assigned to genomic features using STAR^3^ and featureCounts^4^, respectively. We used svaseq R^5^ package (version 3.36.0) to find and remove hidden batch effects. Differential gene expression analysis was performed with DESeq2^6^ R package (version 1.28.1) according to standard protocol.

### ChIP Sequencing

ChIP DNAseq was performed on the Illumina HiSeq 2500 (50 bp, single end) as described in Frank-Bertoncelj et al.^2^. Briefly, ChIP assays were performed using 1 million FLS (Supplementary Table 2) per IP and the following antibodies (all Diagenode): H3K4me3 (0.5 μg, C15411003), H3K27me3 (1 μg, C15410195), H3K27ac (1 μg, C15410196), H3K4me1 (1 μg, C15410194), H3K36me3 (1 μg, C15410192) and H3K9me3 (1 μg, C15410193). The reads were mapped to the GRCh38 human genome reference using Bowtie2^7^ with default settings. The mapped alignment files were further QC’ed with Picard Tools (Broad-Institute, available at: http://broadinstitute.github.io/picard/) to check for duplication rates, unique mapping reads and library complexity. The duplicated reads and non-unique mapping reads were then removed prior to analysis with Picard Tools.

### ChromHMM chromatin state inference

The de-duplicated, uniquely mapping reads of the ChIP sequencing were binarized with the BinarizeBam script provided by the chromHMM software^8^. This script splits the genome into 200bp bins, and compares the coverage of the alignment file at each bin with the input sequence file to determine if any histone modification is present in the bin (1=yes, 0=no). The pre-trained 18 state chromHMM model based on the six histone marks was applied to the binarized bed files, using the MakeSegmentation script provided and the model parameters downloaded from the Roadmap Epigenomics web portal. The methods employed by Ernst et al.^8^ were replicated where possible from the data processing stages to the chromatin state inference.

### ATAC sequencing

Cultured RA FLS were stimulated with 10 ng/ml TNF for 24 h or were left untreated (Supplementary Table 2). 50.000 cells were prepared according to the protocol by Buenrostro et al^9^. ATAC-seq libraries were sequenced on Illumina HiSeq 4000 with 75 bp paired end reads. The reads were QC’d with FastQC for read quality, and the Nextera-transposase adaptors were trimmed with cutadapt^10^. The reads were aligned with Bowtie 2 to the GRCh38 human reference. PCR duplicates were identified and removed by Picard Tools prior to peak calling using MACS2^11^. Both broad and narrow peaks were called as ATAC-seq can have properties of both.

### HiC and capture HiC

Cultured human FLS from RA patients were treated with 10 ng/ml TNF for 24 h or were left untreated (Supplementary Table 2). Cells (1-3×10^7^ per condition), were scratched in 10ml DMEM, spun down, suspended in 35ml DMEM and fixed (2% formaldehyde in DMEM, 10min, RT, with mixing on a rocker). The reaction was quenched with cold 0.125M glycine. Cells were incubated at RT for 5 min, followed by 15 min incubation on ice and centrifugation (1500 rpm, 10 min, 4 °C). Pellets were suspended and washed in cold PBS (1500 rpm, 10 min, 4 °C). Washed pellets were snap frozen and stored at −80 °C. Hi-C libraries from RA FLS samples, were generated as previously described^12^. They were sequenced on Illumina HiSeq 4000 with 75 bp paired end reads. The reads were processed using the HiC Pro pipeline^13^, the correlation between samples were calculated with HiCrep^14^. TADs were called with TADCompare^15^. The regions targeted by the capture HiC (CHiC) were generated based on the LD regions of the lead disease associated SNPs for RA, Juvenile Idiopathic Arthritis (JIA), Psoriatic arthritis (PsA) and Psoriasis (Ps). This resulted in a total of 242 distinct risk variants.120bp capture baits were designed for all HindIII digestion fragments overlapping these regions as previously described in Martin et al.^16^. Significant CHiC interactions were identified through the CHiCAGO pipeline^17^, where the suggested threshold of CHiCAGO score >5 was used. Differential interactions were identified with DESeq2, where the read counts of each interaction were treated similar to the gene count of RNA-seq.

### Transcription factor binding site prediction

We extracted differentially interacting regions from our CHiC data, where the strength in chromatin interaction (log-fold change of read counts between basal and stimulated) correlated with nearby genes. We overlapped these interaction regions (bait and prey fragments) with our ATAC-seq peaks. These ATAC seq peaks were standardised and re-centered to 200bp each.

We then used the findMotifsGenome.pl software from the HOMER suite^18^ to identify significantly enriched motifs in these ATAC-peaks.

### Partitioned Heritability

We defined active chromatin regions of the genome for each FLS sample and publicly available Roadmap samples, based on the union of H3K27ac, H3K4me1, and H3K4me3 histone peaks. We used the partitioned heritability software from the LDSC^19^ package to quantify the non-HLA RA heritability attributed to these active regions in each sample, based on the summary statistics from the Okada et al trans-ethnic meta-analysis^20^.

### Derivation of RA credible set SNPs

For each locus, we dissected distinct RA association signals using approximate conditioning implemented in GCTA^21^, based on: (i) European ancestry summary statistics from the Okada et al. trans-ethnic meta-analysis^20^; and (ii) a reference panel of European ancestry haplotypes from the 1000 Genomes Project to approximate linkage disequilibrium between SNPs. We identified index SNPs for each distinct signal, at a locus-wide significance threshold of *p*<10^− 5^, using the --cojo-slct option. For each locus with multiple distinct signals, we derived the conditional association summary statistics for each distinct signal by conditioning out the effects of all other index SNPs at the locus using the --cojo-cond option.

For each distinct signal, we first calculated the posterior probability, *π*_*j*_, that the *j*th variant is driving the association, given by

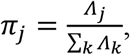

where the summation is over all variants at the locus. In this expression, *Λ*_*j*_ is the approximate Bayes’ factor^22^ for the *j*th variant, given by

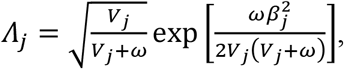

where *β*_*j*_ and *V*_*j*_ denote the estimated allelic effect and corresponding variance from the European ancestry component of Okada et al.^20^. In loci with multiple distinct signals of association, summary statistics were obtained from the approximate conditional analysis. In loci with a single association signal, summary statistics were obtained from unconditional analysis. The parameter *ω* denotes the prior variance in allelic effects, taken here to be 0.04. The 99% credible set for each signal was then constructed by: (i) ranking all variants according to their Bayes’ factor, *Λ*_*j*_; and (ii) including ranked variants until their cumulative posterior probability of driving the association attained or exceeded 0.99.

### Pathway analysis and protein-protein interaction network

The genes assigned to FLS (Supplementary Table 5, column R) and listed in Supplementary Table 5, columns U-Z were analysed by STRINGv11 (interactions settings to medium confidence levels)^23^, ToppFun on ToppGene Suite^24^ and DAVID v6.8^25,26^ with default settings.

### Guide RNA design and cloning

Guide (g)RNAs, targeting the putative upstream *RBPJ* enhancer (locus 23, Supplementary Table 5), were designed using the CRISPOR tool^27^ in the DNA region chr4:26106475-26106675 (hg38) comprising 100bp upstream and 100bp downstream of the RA risk SNP rs874040. Complementary gRNA oligo pairs with 5’ CACC (fwd) and 5’CAAA (rev) overhangs (Microsynth, 100 mM) were phosphorylated and annealed in a termocycler (37°C, 30 min; 95°C 5 min, ramp down to 25°C at 5°C/min using T4PNK (NEB) and 10x T4 ligation buffer (NEB). LentiGuide-puro plasmid, a gift from Feng Zhang (Addgene μplasmid # 52963; http://n2t.net/addgene:52963; RRID:Addgene_52963)^28^, was digested with FastDigest BBsI, Fast AP, 10x Fast Digest Buffer at 37°C, 30 min (Fermentas) followed by the ligation of the annealed gRNA duplex o/n using Quick Ligase (NEB) and 2x Quick Ligation buffer (NEB). One Shot™ Stbl3™ Chemically Competent E. coli (ThermoFisher, C7373-03) were transformed with gRNA containing lentiGuide-Puro plasmids by heat-shocking (45 sec, 42°C) according to manufacturer’s instructions. Plasmid DNA from selected colonies was isolated using QIAprep Spin Miniprep Kit (Qiagen) and Sanger sequenced to confirm the insertion and sequences of cloned gRNAs. To prepare gRNA-containing lentiviral particles, HEK293T cells were transfected with psPAX2, pMD2.G gRNA-containing plasmids, (total 10 μg plasmid DNA, mass ratio 2:1:4, respectively). psPAX2 (Addgene plasmid # 12260; http://n2t.net/addgene:12260; RRID: Addgene_12260) and pMD2.G (Addgene plasmid # 12259; http://n2t.net/addgene:12259; RRID: Addgene_12259) were a gift from Didier Trono. Viral particles were precipitated from the supernatants of transfected HEK293T (24h and 48h) using PEG-itTM Virus Precipitation Solution (5x) according to manufacturer’s protocol (System Biosciences), resuspended in PBS, and stored at −70°C.

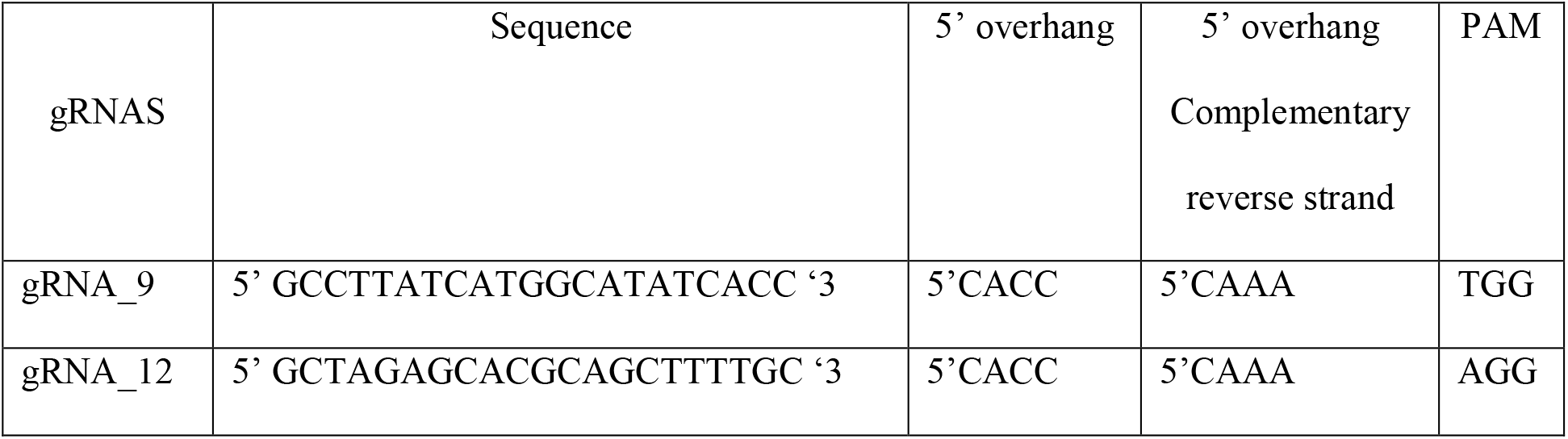

### Activation of enhancer regions with dCas9-VPR

FLS were transduced with Edit-R Lentiviral dCas9-VPR lentiviral particles (hEF1α promoter, Dharmacon). Edit-R Lentiviral dCas9-VPR is a CRISPR activation system, in which a nuclease-deactivated *S. pyogenes* Cas9 (dCas9) is fused to VP64, p65 and Rta transcriptional activators. Stable populations of dCas9-VPR FLS were blasticidin selected (7.5 μg/ml, Horizon) and subsequently transduced with gRNA-containing lentiviral particles. Stable gRNA dCas9-VPR FLS were puromycin selected (5 μg/ml Sigma), lysed, RNA was isolated, followed by reverse transcription and SYBR Green real time PCR as described above. Gene expression was normalized to the average expression of *B2M* (Primer sequence Fwd 5’ AAGCAGCATCATGGAGGTTTG’3, Rev 5’ AAGCAAGCAAGCAGAATTTGGA’3) and *RPLP0* housekeeper genes. Transduction of dCas9-VPR without guide RNA had no effect on *RBPJ* expression.

### Pyrosequencing

DNA from FLS was isolated using the QIAamp DNA Blood kit (Qiagen). DNA regions containing rs874040 (*RBPJ*) were amplified by PCR (Primers: FWD: AGT GTG GAT TGT AGC AGA TAT GTC; REV: biotin-ACC AAG GCA GCC ACA GAA TC; GCT CGG ATG GGG TAT TTC TAG). SNPs were genotyped by pyrosequencing using PyroMark Q48 Advanced Reagents and the PyroMark Q48 Autoprep (both Qiagen) according to manufacturer’s instructions.

### Quantitative Real-time PCR

Total RNA was isolated using the RNeasy Mini Kit (Qiagen) and the Quick RNA MicroPrep Kit (Zymo Research) including on column DNaseI digest and was reverse transcribed. SYBRgreen Real-time PCR was performed (primers: *RBPJ* FWD: GGC TGC AGT CTC CAC GTA CGT C, REV: CTC ACC AAA TTT CCC AGG CGA TGG; *HES1* FWD: ATG GAG AAA AGA CGA AGA GCA AG; REV: TGC CGC GAG CTA TCT TTC TT), including controls (samples containing the untranscribed RNA, dissociation curves). Data were analysed with the comparative CT methods and presented as ΔCT or 2−ΔΔCT as described elsewhere^29^ using *RPLP0* as a housekeeping gene for sample normalization (FWD: GCG TCC TCG TGG AAG TGA CAT CG, REV: TCA GGG ATT GCC ACG CAG GG).

### Cap Analysis Gene Expression (CAGE)

Cultured human FLS from RA patients were treated with 10 ng/ml TNF, 24h or were left untreated (Supplementary Table 2). RNA was isolated using the Quick RNA MicroPrep Kit (Zymo Research). CAGE libraries were prepared and sequenced as previously described in detail^30^. Mapping and identification of CAGE transcription start sites (CTSSs) were performed by DNAFORM (Yokohama, Kanagawa, Japan). In brief, the sequenced CAGE tags were mapped to hg19 using BWA software and HISAT2 after discarding ribosomal RNAs. Identification of CTSSs was performed with the Bioconductor package CAGEr (version 1.16.0)^31^. TSS and enhancer candidate identification and quantification were performed with the Bioconductor package CAGEfightR (version 1.6.0)^32^ with default settings.

## Data availability

The following publicly available datasets were used in this study:

RNA sequencing data (Figs. 2d, 3b): ENA PRJEB14422

Capture HiC data from GM12878 and Jurkat cell lines: NCBI Gene Expression Omnibus

(GEO) GSE69600

All data are available from the authors upon request.

## Notes

### Competing Interest Statement

The authors have declared no competing interest.

### Author Declarations

The studies were approved by the local ethic committees of the University Hospital Zurich, Switzerland and the University of Birmingham, UK

