## Supplemental Figures and Datasets for "Functional genomics atlas of synovial fibroblasts defining rheumatoid arthritis heritability"

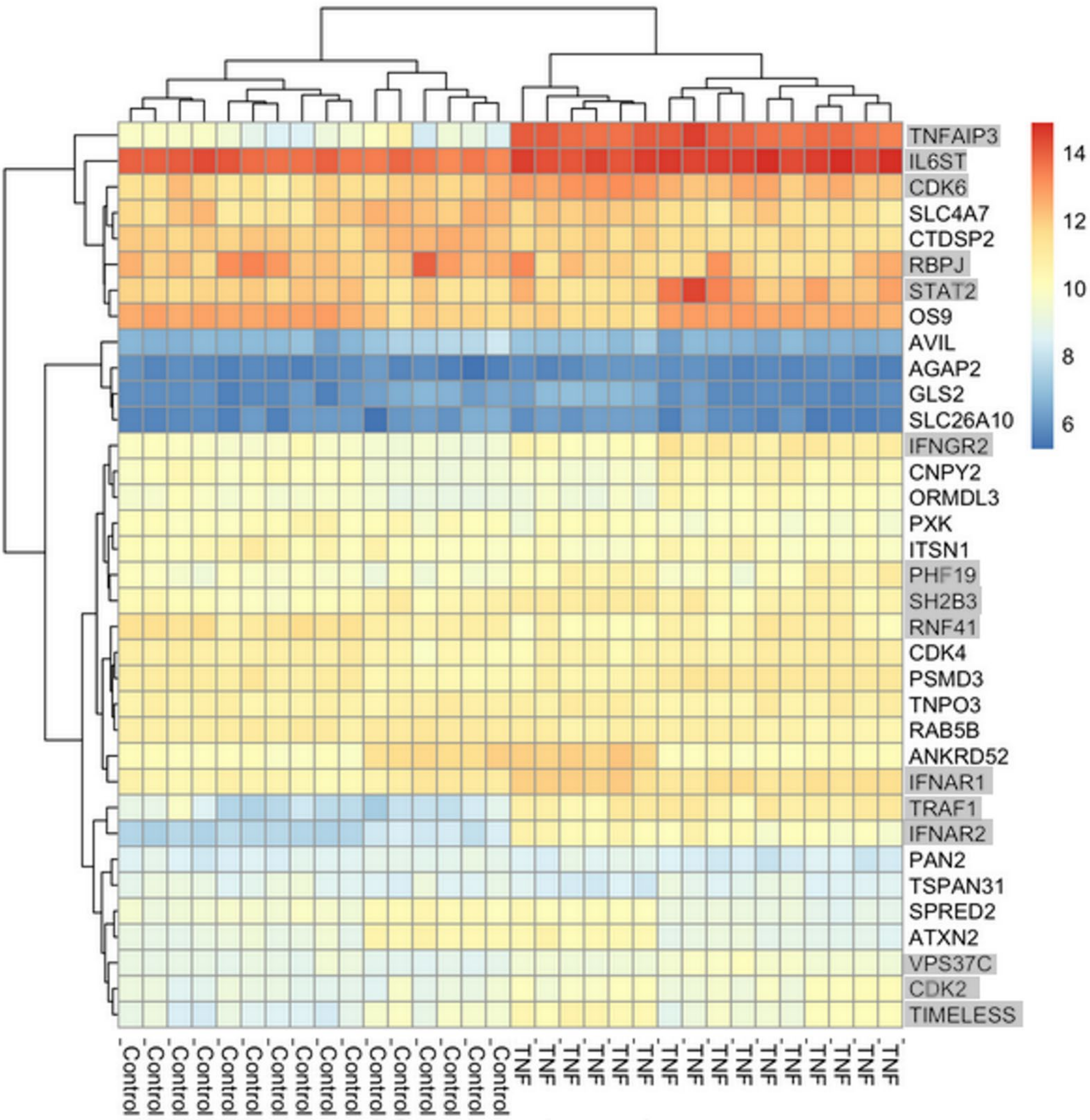


**Supplementary Figure 1.** Heatmap of expression of genes linked to RA loci with a change in chromatin interactions under stimulation with TNF. Genes with an adjusted p-value <0.05 and a log2 fold change >±0.5 are marked in grey.

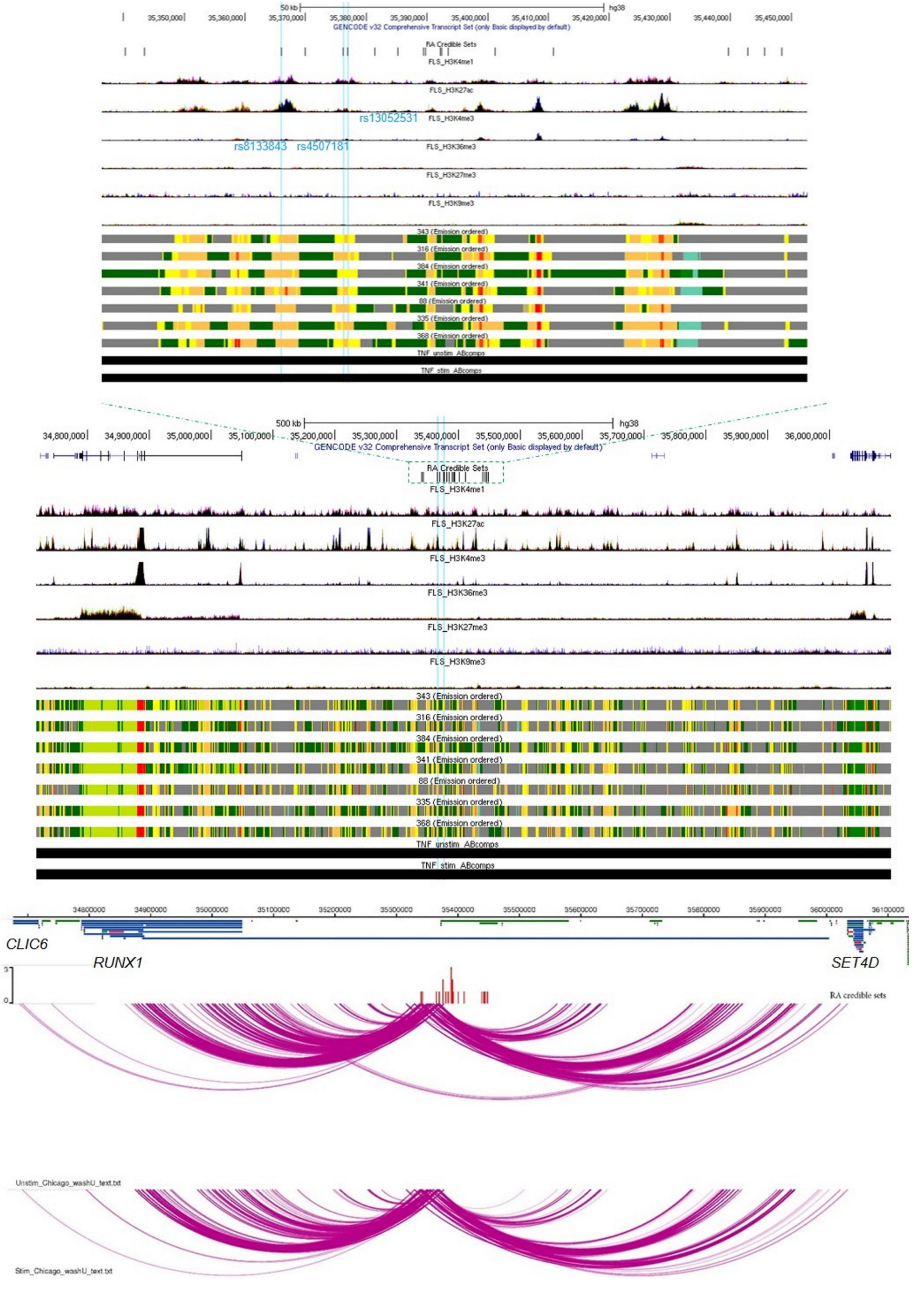


**Supplementary Figure 2.** Fine mapping, epigenetic landscape and chromatin architecture at rs8133843 as an exemplary category 1 locus containing RA credible SNPs. The likely causal credible SNPs are marked with blue lines.

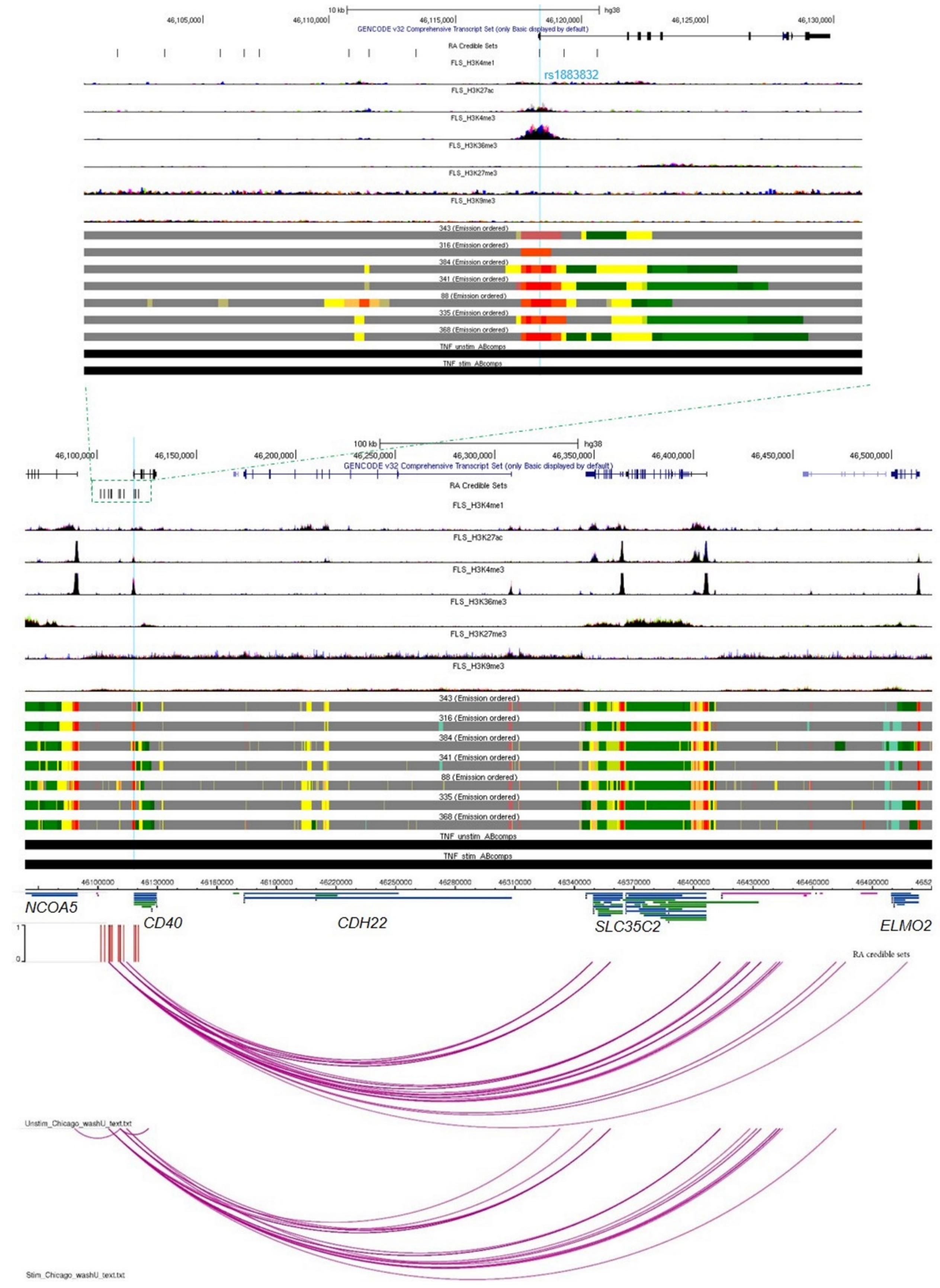


**Supplementary Figure 3.** Fine mapping, epigenetic landscape and chromatin architecture at rs4239702 as an exemplary category 2 locus containing RA credible SNPs. The likely causal credible SNPs are marked with blue lines.

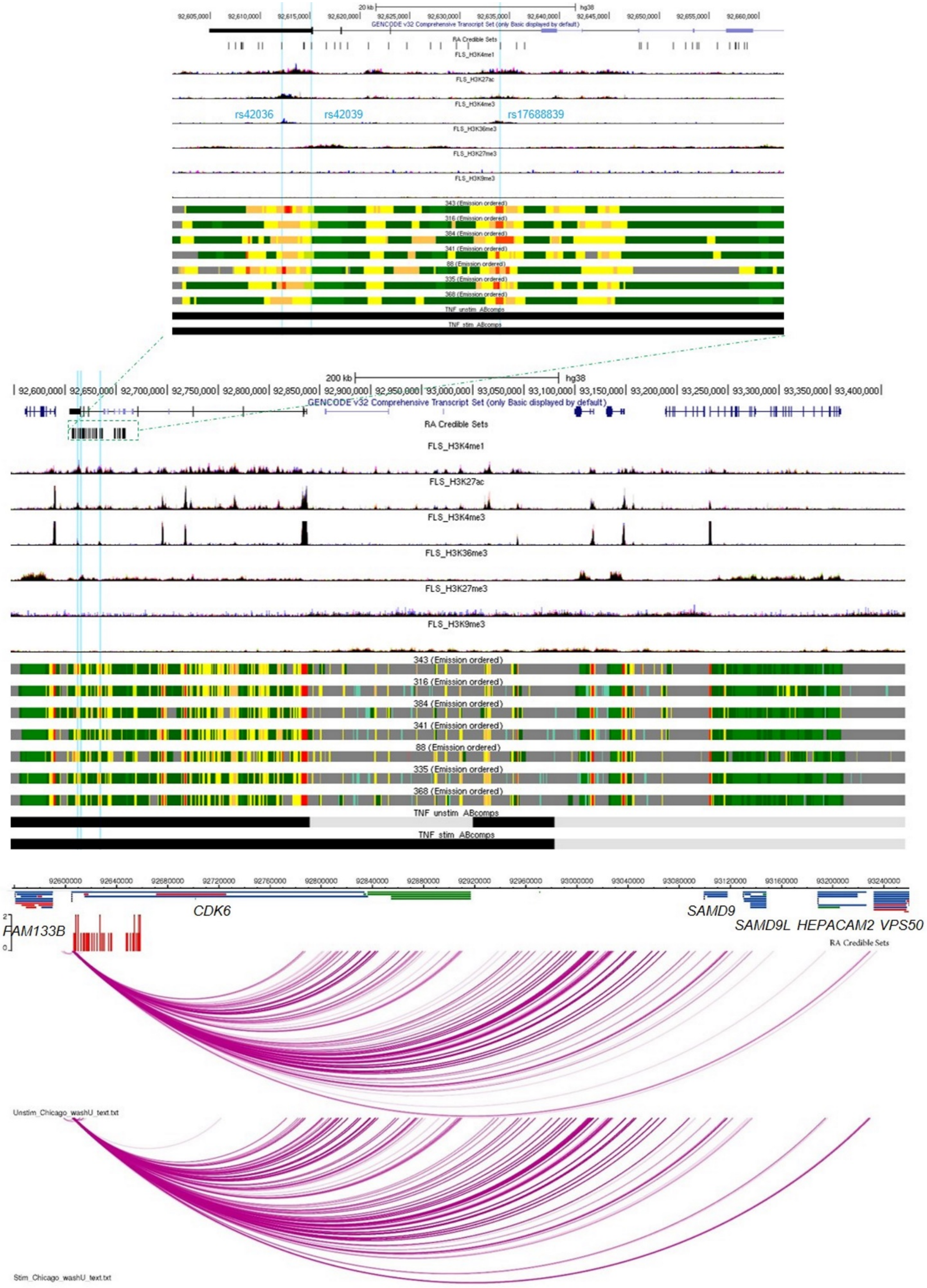


**Supplementary Figure 4.** Fine mapping, epigenetic landscape and chromatin architecture at rs4272 as an exemplary category 3 locus containing RA credible SNPs. The likely causal credible SNPs are marked with blue lines.

**Supplementary Dataset 1.**

**a) Quality check results of the ChIP-seq dataset.**

| | H3K4me1 | | | | H3K4me3 | | | | H3K9me3 | | | |
|---|---|---|---|---|---|---|---|---|---|---|---|---|
| **Sample ID** | **Total Sequences (M)** | **Unmapped reads (M)** | **PERCENT DUPLICATION (%)** | **VALID READS (M)** | **Total Sequences (M)** | **Unmapped reads (M)** | **PERCENT DUPLICATION (%)** | **VALID READS (M)** | **Total Sequences (M)** | **Unmapped reads (M)** | **PERCENT DUPLICATION (%)** | **VALID READS (M)** |
| **88** | 32.8 | 0.5 | 43.8 | 18.2 | 38.7 | 1.04 | 56.7 | 16.3 | 67.9 | 1.45 | 22.2 | 51.7 |
| **316** | 41.4 | 0.5 | 38.7 | 25.1 | 31.9 | 0.32 | 30.2 | 22 | 87.6 | 1.78 | 25.7 | 63.8 |
| **335** | 48.2 | 0.7 | 49.8 | 23.8 | 29.7 | 0.43 | 33.2 | 19.6 | 74.1 | 2.36 | 40.1 | 43 |
| **341** | 42.2 | 0.6 | 40.6 | 24.7 | 29.4 | 0.56 | 43 | 16.4 | 66.5 | 1.49 | 19.1 | 52.6 |
| **343** | 43.5 | 0.6 | 51.6 | 20.8 | 29.8 | 0.47 | 28.9 | 20.9 | 49.2 | 1.1 | 19.2 | 38.9 |
| **368** | 51.6 | 0.5 | 54.6 | 23.2 | 37.7 | 0.61 | 45.2 | 20.3 | 61.4 | 1.52 | 43.3 | 34 |
| **384** | 37.4 | 0.8 | 62 | 13.9 | 30.1 | 0.42 | 32 | 20.2 | 59.3 | 1.24 | 22 | 45.3 |

| | H3K27ac | | | | H3K27me3 | | | | H3K36me3 | | | |
|---|---|---|---|---|---|---|---|---|---|---|---|---|
| **Samples** | **Total Sequences (M)** | **Unmapped reads (M)** | **PERCENT DUPLICATION (%)** | **VALID READS (M)** | **Total Sequences (M)** | **Unmapped reads (M)** | **PERCENT DUPLICATION (%)** | **VALID READS (M)** | **Total Sequences (M)** | **Unmapped reads (M)** | **PERCENT DUPLICATION (%)** | **VALID READS (M)** |
| **88** | 40.4 | 0.5 | 37.9 | 24.8 | 48.4 | 2.81 | 78.4 | 9.8 | 62.5 | 0.55 | 15.2 | 52.5 |
| **316** | 39.7 | 0.39 | 28.4 | 28.1 | 53.1 | 0.69 | 52.2 | 25.1 | 64.4 | 0.53 | 18.9 | 51.8 |
| **335** | 44.6 | 0.83 | 52.3 | 20.9 | 51.3 | 0.76 | 64.9 | 17.7 | 84.2 | 1 | 21.8 | 65.1 |
| **341** | 35.8 | 0.62 | 54.9 | 15.9 | 52.6 | 0.71 | 52.6 | 24.6 | 70.2 | 0.97 | 22.6 | 53.6 |
| **343** | 36.9 | 0.51 | 44.5 | 20.2 | 52 | 0.87 | 58.8 | 21.1 | 50 | 0.61 | 18 | 40.5 |
| **368** | 45.5 | 0.91 | 52.6 | 21.1 | 51.5 | 0.74 | 56.7 | 22 | 63.4 | 0.87 | 28.7 | 44.6 |
| **384** | 47.1 | 1.5 | 80.1 | 9.1 | 49.3 | 0.86 | 59.5 | 19.6 | 68.3 | 0.63 | 16.3 | 56.6 |

**b) Hi-C read information and quality checks.**
Samples are unstimulated controls, or have been TNF stimulated (_TNF rows).

| Sample ID | Joint | Total Reads (M) | | Unique Alignments (M) | | Paired Reads (M) | Valid Pairs (M) | Percentage Mapped (%) |
|---|---|---|---|---|---|---|---|---|
| | | R1 | R2 | R1 | R2 | | | |
| **SF_412** | Wrist | 103 | 103 | 80 | 79 | 63 | 51 | 61.85 |
| **SF_412_TNF** | Wrist | 110 | 110 | 88 | 87 | 71 | 61 | 64.88 |
| **SF_415A** | Wrist | 112 | 112 | 84 | 83 | 64 | 48 | 57.2 |
| **SF_415A_TNF** | Wrist | 591 | 591 | 467 | 463 | 371 | 347 | 62.91 |
| **SF_415B** | MCP (Hand) | 56 | 56 | 41 | 41 | 30 | 20 | 54.31 |
| **SF_415B_TNF** | MCP (Hand) | 96 | 96 | 76 | 75 | 60 | 51 | 62.16 |
| **SF_420** | Shoulder | 120 | 120 | 94 | 92 | 72 | 63 | 60.64 |
| **SF_420_TNF** | Shoulder | 291 | 291 | 230 | 222 | 174 | 154 | 59.95 |
| **SF_424** | MCP (Hand) | 79 | 79 | 63 | 62 | 51 | 46 | 64.63 |
| **SF_424_TNF** | MCP (Hand) | 83 | 83 | 67 | 66 | 54 | 48 | 64.57 |
| **SF_427** | Knee | 419 | 419 | 321 | 316 | 249 | 170 | 59.39 |
| **SF_427_TNF** | Knee | 277 | 277 | 213 | 209 | 166 | 115 | 60.1 |
| **SF_429** | Shoulder | 291 | 291 | 236 | 233 | 193 | 164 | 66.26 |
| **SF_429_TNF** | Shoulder | 301 | 301 | 245 | 241 | 201 | 163 | 66.95 |

**c) Capture Hi-C read information and quality checks.**
Samples are unstimulated controls or have been TNF stimulated (_TNF rows).

| Sample ID (capture Hi-C) | Joint | Total Reads (M) | | Unique Alignments (M) | | Paired Read (M) | Valid Pairs (M) | Percentage Mapped (%) |
|---|---|---|---|---|---|---|---|---|
| | | R1 | R2 | R1 | R2 | | | |
| **SF_412** | Wrist | 203 | 203 | 158 | 155 | 125 | 104 | 61.8 |
| **SF_412_TNF** | Wrist | 332 | 332 | 266 | 262 | 218 | 189 | 65.73 |
| **SF_415A** | Wrist | 247 | 247 | 187 | 186 | 143 | 124 | 57.93 |
| **SF_415A_TNF** | Wrist | 155 | 155 | 123 | 122 | 100 | 94 | 64.55 |
| **SF_415B** | MCP (Hand) | 336 | 336 | 239 | 237 | 173 | 122 | 51.7 |
| **SF_415B_TNF** | MCP (Hand) | 310 | 310 | 250 | 247 | 200 | 172 | 64.78 |
| **SF_420** | Shoulder | 217 | 217 | 177 | 175 | 143 | 127 | 65.98 |
| **SF_420_TNF** | Shoulder | 156 | 156 | 128 | 126 | 103 | 92 | 66.05 |
| **SF_424** | MCP (Hand) | 173 | 173 | 144 | 141 | 117 | 105 | 67.52 |
| **SF_424_TNF** | MCP (Hand) | 154 | 154 | 126 | 125 | 104 | 95 | 67.51 |
| **SF_427** | Knee | 218 | 218 | 161 | 159 | 124 | 92 | 56.84 |
| **SF_427_TNF** | Knee | 216 | 216 | 167 | 164 | 131 | 99 | 60.6 |
| **SF_429** | Shoulder | 216 | 216 | 175 | 173 | 145 | 124 | 67.11 |
| **SF_429_TNF** | Shoulder | 252 | 252 | 206 | 202 | 171 | 141 | 68.04 |

**d) ATAC-seq read information and quality checks.**
Samples are unstimulated controls, or have been TNF stimulated (_TNF rows).

| Sample ID (ATAC) | READ PAIRS EXAMINED (M) | READ PAIR DUPLICATES (M) | PERCENT DUPLICATION (%) | ESTIMATED LIBRARY SIZE (M) |
|---|---|---|---|---|
| **SF_346** | 94.3 | 68.5 | 72.7 | 26.5 |
| **SF_346_TNF** | 85.3 | 37.7 | 44.1 | 66.4 |
| **SF_415B** | 75.1 | 47.9 | 63.8 | 29.6 |
| **SF_415B_TNF** | 73.7 | 27 | 36.7 | 75.5 |
| **SF_420** | 78.9 | 57.1 | 72.3 | 22.5 |
| **SF_420_TNF** | 67.3 | 54 | 80.3 | 13.3 |
| **SF_424** | 72.8 | 59.2 | 81.3 | 13.7 |
| **SF_424_TNF** | 90.1 | 59.3 | 65.8 | 33.1 |
| **SF_427** | 81.6 | 55.1 | 67.5 | 28.1 |
| **SF_427_TNF** | 63.3 | 42.7 | 67.5 | 21.8 |
| **SF_429** | 95.3 | 59.9 | 62.8 | 38.8 |
| **SF_429_TNF** | 65.8 | 32.7 | 49.7 | 41.9 |

**e) RNA-seq data quality and alignment summary.**

Samples are unstimulated controls or have been TNF stimulated (_TNF rows).

| Sample ID | Total bases (M) | Aligned bases (M) | Coding bases (%) | UTR bases (%) | Intronic bases (%) | Intergenic bases (%) | Usable bases (%) |
|---|---|---|---|---|---|---|---|
| **SF_292** | 10092.8 | 9943.9 | 16.7 | 18.8 | 27.3 | 37.2 | 35 |
| **SF_292_TNF** | 7794.8 | 7673.9 | 10.6 | 16.2 | 27.2 | 46.1 | 26.3 |
| **SF_336** | 8206.8 | 8072 | 10.4 | 20.3 | 25.5 | 43.8 | 30.2 |
| **SF_336_TNF** | 7983.2 | 7846.4 | 12.6 | 18.8 | 26.6 | 42 | 30.9 |
| **SF_346** | 7263.5 | 7130.7 | 9.9 | 16.2 | 27.3 | 46.6 | 25.6 |
| **SF_346_TNF** | 6830.9 | 6705.5 | 11.3 | 18.4 | 26.6 | 43.6 | 29.2 |
| **SF_407** | 6360.5 | 6251 | 20.4 | 20.5 | 24.5 | 34.6 | 40.2 |
| **SF_407_TNF** | 8051.5 | 7954.8 | 6.7 | 17.1 | 28.1 | 48.1 | 23.5 |
| **SF_415B** | 4927.7 | 4858.9 | 6 | 16.2 | 27.6 | 50.2 | 22 |
| **SF_415B_TNF** | 8369.4 | 8271.4 | 6.2 | 17.1 | 28.8 | 47.9 | 22.9 |
| **SF_435** | 8165.7 | 8052.1 | 5.2 | 15.9 | 27 | 52 | 20.7 |
| **SF_435_TNF** | 10682.4 | 10529.9 | 6 | 17 | 29.2 | 47.7 | 22.7 |

**f) CAGE-seq data quality and alignment summary.**

Samples are unstimulated controls or have been TNF stimulated (_TNF rows).

| Sample ID | Total Reads (M) | rRNA-Filtered (M) | BWA-Mapped (M) | HISAT2-Mapped (M) | Total Mapped (M) | Percentage Mapped (%) |
|---|---|---|---|---|---|---|
| **SF_276** | 25.8 | 8.7 | 14.3 | 1.7 | 16.0 | 62.1 |
| SF_276_TNF | 25.4 | 7.6 | 15.2 | 1.4 | 16.6 | 65.5 |
| **SF_415** | 24.9 | 4.7 | 17.7 | 1.6 | 19.3 | 77.6 |
| **SF_415_TNF** | 20.7 | 4.3 | 13.9 | 1.4 | 15.3 | 74.2 |
| **SF_420** | 25.0 | 6.2 | 16.0 | 1.5 | 17.4 | 69.8 |
| SF_420_TNF | 25.3 | 5.6 | 17.0 | 1.5 | 18.5 | 73.0 |
| SF_424 | 24.9 | 7.6 | 14.6 | 1.7 | 16.4 | 65.8 |
| SF_424_TNF | 27.5 | 14.2 | 10.6 | 1.6 | 12.2 | 44.5 |
| **SF_427** | 24.3 | 7.5 | 14.1 | 1.6 | 15.7 | 64.6 |
| **SF_427_TNF** | 23.6 | 5.9 | 15.2 | 1.4 | 16.6 | 70.2 |
| **SF_429** | 25.6 | 7.7 | 15.0 | 1.8 | 16.8 | 65.8 |
| **SF_429_TNF** | 25.0 | 5.2 | 17.3 | 1.6 | 18.9 | 75.5 |
| **SF_460** | 20.8 | 4.8 | 13.8 | 1.4 | 15.2 | 73.1 |
| **SF_460_TNF** | 24.0 | 4.7 | 16.5 | 1.6 | 18.1 | 75.6 |

BWA = Burrows-Wheeler Aligner

**Supplementary Dataset 2.** Known motif enrichment analysis using HOMER at enhancer sites with open chromatin (as defined by ATAC-seq).

STIMULATED ATAC-seq peaks cut-off p-value = 1.00E-04

**CHiC up/RNA up**

| Rank | Motif | Name | P-value | log P-pvalue | q-value (Benjamini) | # Target Sequences with Motif | % of Targets Sequences with Motif | # Back-ground Sequences with Motif | % of Back-ground Sequences with Motif |
|---|---|---|---|---|---|---|---|---|---|
| 1 | 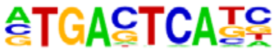 | AP-1(bZIP)/ThioMac-PU.1-ChIP-Seq(GSE21512)/Homer | 1.00E-12 | -2.97E+01 | 0 | 45 | 20.83% | 2901.5 | 5.90% |
| 2 | 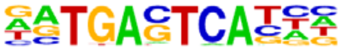 | Atf3(bZIP)/GBM-ATF3-ChIP-Seq(GSE33912)/Homer | 1.00E-10 | -2.35E+01 | 0 | 38 | 17.59% | 2576.3 | 5.24% |
| 3 | 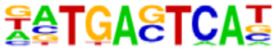 | BATF(bZIP)/Th17-BATF-ChIP-Seq(GSE39756)/Homer | 1.00E-08 | -1.91E+01 | 0 | 34 | 15.74% | 2494.7 | 5.07% |
| 4 | 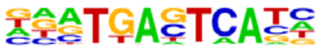 | Fra1(bZIP)/BT549-Fra1-ChIP-Seq(GSE46166)/Homer | 1.00E-06 | -1.58E+01 | 0 | 28 | 12.96% | 2061.2 | 4.19% |
| 5 | 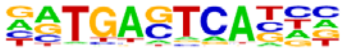 | Fosl2(bZIP)/3T3L1-Fosl2-ChIP-Seq(GSE56872)/Homer | 1.00E-04 | -1.10E+01 | 0.0011 | 18 | 8.33% | 1274.9 | 2.59% |

**CHiC up/RNA down**

| Rank | Motif | Name | P-value | log P-pvalue | q-value (Benjamini) | # Target Sequences with Motif | % of Targets Sequences with Motif | # Back-ground Sequences with Motif | % of Back-ground Sequences with Motif |
|---|---|---|---|---|---|---|---|---|---|
| 1 | 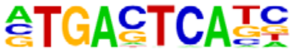 | AP-1(bZIP)/ThioMac-PU.1-ChIP-Seq(GSE21512)/Homer | 1.00E-24 | -5.62E+01 | 0 | 76 | 23.31% | 2904.8 | 5.89% |
| 2 | 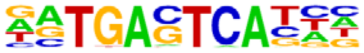 | Atf3(bZIP)/GBM-ATF3-ChIP-Seq(GSE33912)/Homer | 1.00E-21 | -5.02E+01 | 0 | 67 | 20.55% | 2511.4 | 5.09% |
| 3 | 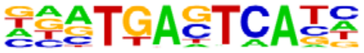 | Fra1(bZIP)/BT549-Fra1-ChIP-Seq(GSE46166)/Homer | 1.00E-19 | -4.52E+01 | 0 | 57 | 17.48% | 2008.9 | 4.07% |
| 4 | 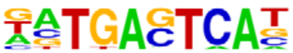 | BATF(bZIP)/Th17-BATF-ChIP-Seq(GSE39756)/Homer | 1.00E-17 | -4.02E+01 | 0 | 60 | 18.40% | 2473.5 | 5.01% |
| 5 | 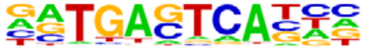 | Fosl2(bZIP)/3T3L1-Fosl2-ChIP-Seq(GSE56872)/Homer | 1.00E-12 | -2.82E+01 | 0 | 34 | 10.43% | 1147.5 | 2.32% |
| 6 | 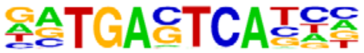 | Jun-AP1(bZIP)/K562-cJun-ChIP-Seq(GSE31477)/Homer | 1.00E-09 | -2.25E+01 | 0 | 26 | 7.98% | 846.1 | 1.71% |
| 7 | 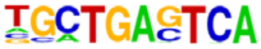 | Bach2(bZIP)/OCILy7-Bach2-ChIP-Seq(GSE44420)/Homer | 1.00E-04 | -1.12E+01 | 0.0006 | 16 | 4.91% | 668.4 | 1.35% |
| 8 | 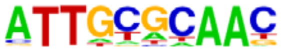 | CEBP(bZIP)/ThioMac-CEBPb-ChIP-Seq(GSE21512)/Homer | 1.00E-04 | -9.37E+00 | 0.0034 | 40 | 12.27% | 3191.3 | 6.47% |
